# TACR3 variant confers resilience to aging and Alzheimer’s disease

**DOI:** 10.64898/2026.06.06.26355071

**Authors:** Nicolas Ruffini, Florian Udo Fischer, Robert Subirana Slotos, Julia Goschke, Leon Scholz, Kristel Knaepen, Stefan Hüttelmaier, Helen Morrison, Theresa Steffan, Ann-Sophie Pabst, Jennifer Winter, Bernhard Baier, Andreas Mierau, Harald Binder, Alexander Drzezga, Stefan Teipel, Andreas Fellgiebel, Kristina Endres, Oliver Tüscher, the Alzheimer’s Disease Neuroimaging Initiative, the German AgeGain study group

## Abstract

**Background:** While genetic factors strongly influence brain aging trajectories, variants conferring cognitive resilience remain poorly characterized. The neurokinin-3 receptor (NK3-R), encoded by Tachykinin Receptor 3 (*TACR3)*, modulates cholinergic signaling in memory circuits vulnerable to aging. Previous studies linked the non-WT expression of the *TACR3* variant rs2765 with cognitive decline and reduced volume of the hippocampus and basal forebrain, but systematic replication and mechanistic validation were lacking.

**Methods:** We investigated rs2765 in the preregistered AgeGain cohort of cognitively healthy older adults (n=188) with independent validation in the ADNI cohort (n=809) which includes persons with and without Alzheimer’s Disease (AD) that show healthy cognition, mild cognitive impairment or dementia. Analyses integrated structural neuroimaging, longitudinal cognitive assessments, epigenetic aging (PhenoAge), genome-wide methylation profiling, and mechanistic validation through luciferase assays and cross-species protein expression studies.

**Results:** The infrequent protective rs2765 WT variant, found in 12.8% of Europeans, conferred 49% slower cognitive decline (*p* = 0.002) for amyloid-positive individuals of the ADNI cohort and 3.7 years younger epigenetic age (*p* = 0.013, 95% CI: 0.79-6.67 years) in the cognitively healthy AgeGain cohort. WT carriers showed larger hippocampal and basal forebrain volumes across cohorts, with Allen Brain Atlas integration revealing these outcomes to occur exclusively in regions where *TACR3* expression positively correlated with gray matter volume. Mechanistically, the non-WT variant ameliorated RBMX-mediated post-transcriptional regulation, reducing NK3-R protein expression by 25-40% *in vitro* and *ex vivo* murine brain slice models. Senescence-accelerated mice exhibited reduced endogenous NK3-R expression, phenocopying the predicted functional consequences of the variant. In AgeGain participants, genome-wide methylation profiling identified 2,313 differentially methylated CpGs affecting 228 pathways spanning glutamatergic signaling, acetylcholine receptor pathways, chromatin remodeling, and angiogenesis, suggesting coordinated molecular reprogramming from synaptic function to systemic aging.

**Conclusions:** rs2765 WT confers resilience to age- and AD-related cognitive decline through RBMX-dependent regulation of NK3-R expression, with effects of remarkable size cascading from memory to systemic aging. rs2765 genotyping could stratify individuals for NK3-R modulator therapy (e.g., fezolinetant or senktides) and identify those maintaining function despite pathological burden, complementing APOE-based risk assessment in precision geromedicine.

## Main

Aging exhibits remarkable heterogeneity across individuals, with some maintaining functional independence and cognitive resilience while others experience rapid multisystem decline (Ferrucci and Kuchel, 2021; Tian *et al*., 2023). While extreme longevity has a strong genetic component, with rare coding variants converging on conserved longevity pathways (Lin *et al*., 2021), the specific variants and mechanisms driving cognitive and brain aging resilience remain poorly characterized, particularly within vulnerable memory systems. Here, we identified rs2765, a common 3’UTR variant of *TACR3* carried by 87% of Europeans that accelerates aging, with the infrequent WT allele conferring resilience through mechanistic pathways validated from molecular to systemic scales.

Memory-related brain networks show pronounced vulnerability to aging, with the basal forebrain emerging as an early indicator of broader neurodegenerative processes: its degeneration, precedes and predicts cortical spread of Alzheimer’s disease (AD) pathology (Schmitz, Nathan Spreng, and Alzheimer’s Disease Neuroimaging Initiative, 2016). The cholinergic modulation of these memory circuits positions them as ideal systems for identifying genetic resilience factors, as basal forebrain volume correlates with general intelligence and enables adaptive network reconfiguration following damage (Wolf *et al*., 2014; Ray *et al*., 2015).

The Tachykinin Receptor 3 (TACR3) gene encodes the Neurokinin 3 Receptor (NK3-R), which modulates neuronal excitability across brain regions critical for memory and cognition potentially through mechanisms like the ’excited or not’ hypothesis, controlling excitability setpoints of different neuronal populations (Zhang, Wang and Chu, 2020). Prior work established *TACR3*’s relevance to cognitive aging: NK3-R agonists enhance learning and memory in aged rats by increasing acetylcholine release 2-4 fold (de Souza Silva *et al*., 2013). In human patients with cognitive impairment, de Souza Silva et al. (2013) found smaller hippocampal volumes in carriers of the rs2765 variant, located in the 3’UTR of *TACR3,* while Teipel et al. (2015) detected steeper age-related basal forebrain volume decline in carriers of the rs2765 variant across 1,967 healthy individuals (Figure 1A).

**Figure 1:**
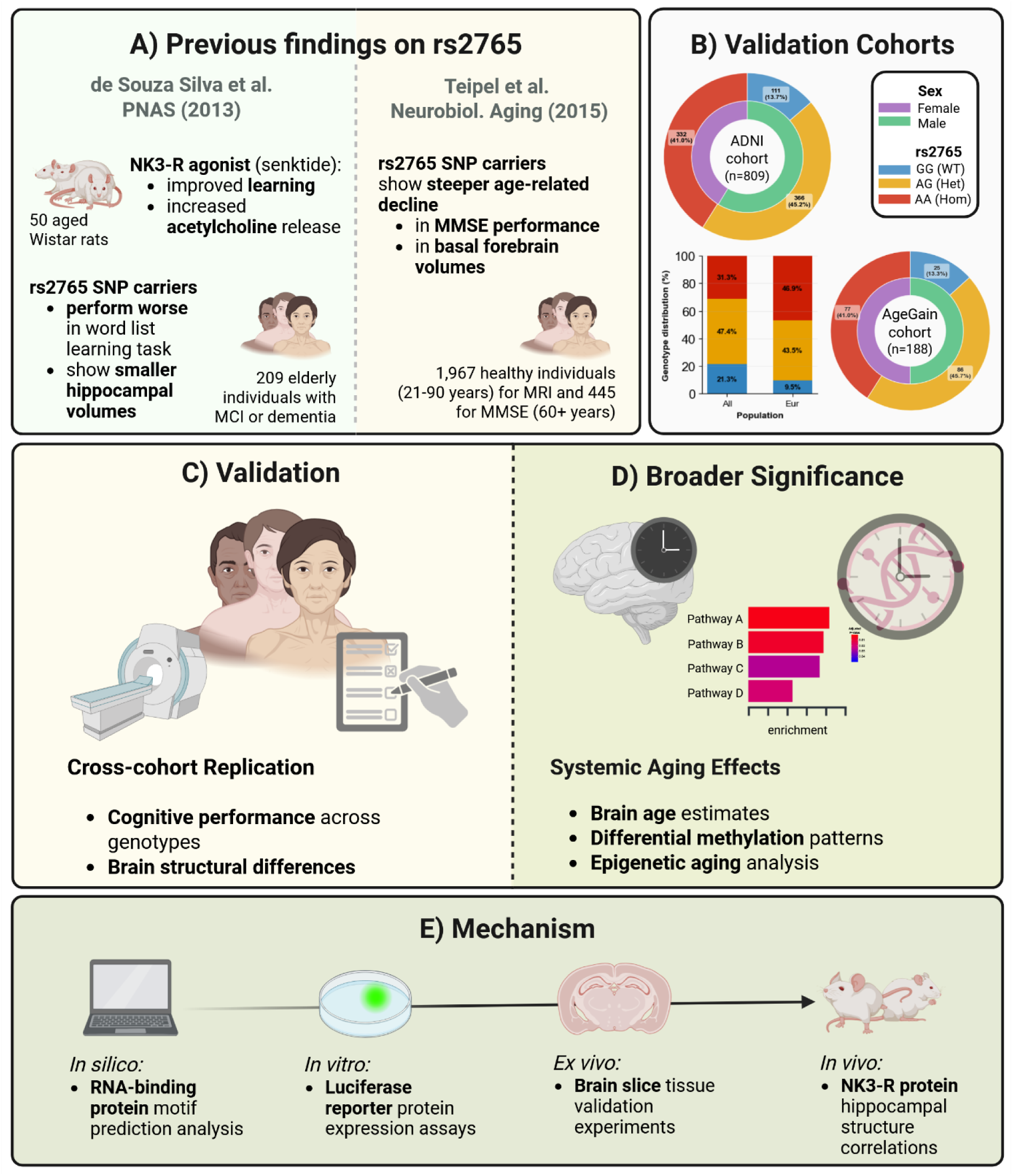
Study design and multi-level validation of rs2765 effects on aging resilience. **A)** Previous literature foundations establishing rs2765 associations with cognitive aging. De Souza Silva et al. (2013) demonstrated NK3-R agonist effects on acetylcholine release and learning in aged rats. In elderly individuals with MCI or dementia, they further showed that rs2765 single nucleotide polymorphism (SNP) carriers, i.e. non-WT carriers, performed worse in word-list learning tasks and had smaller hippocampal volumes. Teipel et al. (2015) reported steeper age-related decline in MMSE scores and basal forebrain volumes in human rs2765 SNP carriers across 1,967 healthy individuals (445 for MMSE analysis). **B)** Our validation approach examining cohort composition and demographic distributions for AgeGain (n=188) and ADNI (n=809; of which n=121 cognitively normal amyloid negative) datasets. Inner rings indicate sex distribution; outer rings show rs2765 genotype frequencies (AA, homozygous variant; AG, heterozygous; GG, WT). Comparison with population reference data shows that the protective GG WT genotype represents a minority across populations globally (African: 16.9%, Asian: 24.9%) and is particularly low with 12.8% in European-ancestry populations (*dbSNP rs2765*, 2025). **C)** Cross-cohort replication approach examining cognitive performance across genotypes and brain structural differences using neuroimaging and longitudinal cognitive assessments. **D)** Assessment of global brain aging (brain age) and systemic aging biomarkers (methylation patterns, PhenoAge) extending beyond regional structural differences. **E)** Mechanistic pathway elucidation spanning *in silico* RNA-binding protein motif prediction analysis, *in vitro* luciferase reporter protein expression assays, *ex vivo* brain slice tissue validation experiments, and *in vivo* NK3-R protein hippocampal structure correlations to establish the molecular basis underlying rs2765 effects on aging resilience. Created in BioRender. Ruffini, N. (2025) https://BioRender.com/i44fgy2

Understanding genetic determinants of aging resilience, i.e. the mechanism that help to maintain function despite accumulating pathology during aging and age-related diseases (Wolf, Fischer, Fellgiebel, and Alzheimer’s Disease Neuroimaging Initiative, 2019; Stern *et al*., 2020), and especially as observed in cognitively healthy centenarians (Rohde *et al*., 2025), requires identifying variants with mechanistic validation spanning molecular to systemic scales. The location of rs2765 in *TACR3*’s 3’UTR suggests post-transcriptional regulation as one underlying mechanism (Romo, Findlay and Burge, 2024).

We hypothesized that rs2765 drives aging resilience through post-transcriptional control of NK3-R protein expression in memory circuits, with cascading effects on systemic aging. To test this, we pursued three complementary lines of investigation (Figure 1). First, we systematically replicated previous rs2765 associations through cross-sectional analyses in our AgeGain cohort of cognitively healthy older adults (Wolf *et al*., 2018; Fischer *et al*., 2025) and independent validation in the longitudinal Alzheimer’s Disease Neuroimaging Initiative (ADNI) cohort, spanning the clinical spectrum from cognitively healthy and normal aging to dementia in persons with and without amyloidosis (Figure 1A-C). In the ADNI cohort, we additionally investigated whether rs2765 moderates longitudinal cognitive outcomes specifically in AD across its clinical spectrum. Second, we investigated whether regional brain-specific effects extended to global brain aging and systemic aging by examining brain age prediction, epigenetic aging (PhenoAge), and genome-wide methylation patterns (Figure 1D). Third, we elucidated the molecular mechanism through *in silico* RNA-binding protein motif analysis, followed by *in vitro* luciferase reporter assays, *ex vivo* brain slice validation, and *in vivo* protein expression studies linking rs2765 to NK3 receptor levels and hippocampal structure (Figure 1E). Our work establishes rs2765 as a mechanistically validated determinant of aging resilience, with immediate implications for NK3-R antagonist safety monitoring, a new stratified indication for NK3-R agonist (e.g. senktides) and precision geromedicine.

## Results

We investigated the effects of rs2765 through three complementary approaches to establish its role in aging resilience: (i) systematic replication across cohorts, (ii) assessment of biological significance through systemic aging markers, and (iii) mechanistic characterization of the underlying pathway. Our analyses utilized the preregistered cognitively healthy AgeGain cohort (n=188 genotyped; 25 WT carriers) and ADNI validation cohort (n=809; 111 WT carriers), with sample sizes varying by data modality availability (detailed in Supplementary Table 1, Supplementary Figure 1).

### Cross-cohort replication establishes rs2765 as a genetic determinant of brain aging resilience

Our regionally masked voxel-based morphometry analysis replicated and extended previous observations (de Souza Silva *et al*., 2013; Teipel *et al*., 2015), revealing that rs2765 WT carriers exhibited significantly larger gray matter volumes (GMV) in both hippocampal and basal forebrain regions in cognitively healthy older adults (AgeGain; Figure 2A, B), effectively bridging these previous observations. This beneficial effect of rs2765 WT on GMV extended to the ADNI cohort (Figure 2C, D) spanning cognitively healthy participants to diagnosed dementia patients with and without amyloidosis. These results were consistent with ROI-based automatic hippocampal segmentation volume analyses, where WT carriers showed bilateral hippocampal volume increases with consistent effect magnitudes across cohorts (AgeGain: β = 0.32 SD, p = 0.039; ADNI: β = 0.23 SD, p = 0.015; Figure 2B, D; Supplementary Table 1).

**Figure 2:**
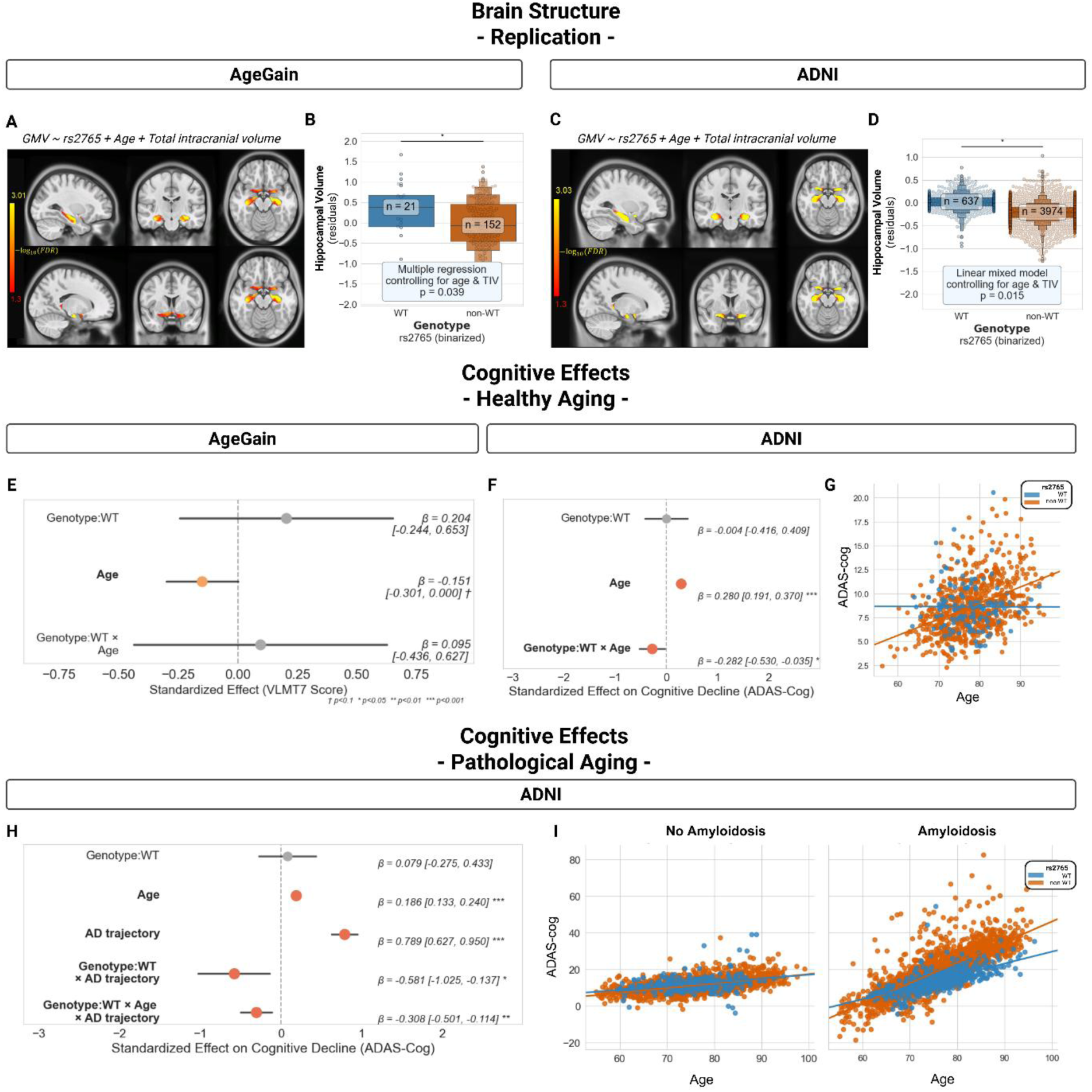
Cross-cohort validation of rs2765 effects on brain structure and cognitive outcomes. **A)** Statistical parametric maps showing voxels of significantly larger GMV in rs2765 WT carriers in the AgeGain cohort, within masked hippocampal and basal forebrain regions of interest (FDR corrected, p < 0.05, N = 173 with MRI of the 188 genotyped). **B)** Hippocampal volumes stratified by rs2765 genotype in AgeGain (N = 173). Values shown are residuals from a multiple regression model controlling for age and total intracranial volume (TIV). WT carriers show significantly larger volumes (multiple regression p = 0.039). **C)** Independent replication in the full ADNI cohort (with and without amyloidosis) showing voxels of significantly larger GMV in rs2765 WT carriers within hippocampal and basal forebrain regions of interest (N = 799). **D)** Hippocampal volumes by rs2765 genotype in ADNI (N = 799). Values shown are residuals from mixed-effects model controlling for age and TIV and per subject intercepts. WT carriers show significantly larger volumes (mixed-effects p = 0.015; 110 WT, 689 non-WT, 4,611 observations plotted in total). **E)** Forest plot of VLMT7 regression model results in AgeGain participants, showing no significant genotype effect (p = 0.37) or genotype×age interaction (p = 0.72), with marginal age main effect (p = 0.050, n=188 subjects). **F)** Forest plots of rs2765 effects on longitudinal cognitive decline (ADAS-Cog, higher values indicate worse performance) in cognitively healthy ADNI participants without amyloidosis, showing significant genotype×age interaction (β = -0.28, p = 0.026, N = 121 subjects, 853 observations) showing complete attenuation of age-related decline in WT carriers. **G)** Partial regression plot showing ADAS-Cog scores (residualized for random intercepts per subject) versus age in cognitively healthy participants, illustrating differential decline trajectories by rs2765 genotype. **H)** Forest plots demonstrating stronger attenuation of cognitive decline for rs2765 WT specifically in participants with amyloid pathology, showing significant three-way interaction (genotype×age×amyloidosis: β = -0.31, p = 0.002, N = 729, 5265 observations) representing complete attenuation of AD-specific cognitive decline acceleration in pathological contexts. **I)** Partial regression plot showing longitudinal cognitive trajectories (residualized for random intercepts per subject) in ADNI participants split by amyloidosis and stratified by rs2765 genotype, demonstrating reduced decline rates in WT carriers.

To determine the functional significance of these structural differences, we examined cognitive decline across genotypes. In the AgeGain cohort, cross-sectional cognitive data did not indicate attenuated decline of verbal learning (VLMT7) with age in rs2765 WT cognitively healthy older adults (age×genotype interaction β = 0.10, p=0.72; Figure 2E). However, in the ADNI cohort, analyses of longitudinal, cognitive data in cognitively healthy amyloid-negative older adults demonstrated attenuated decline in WT-carriers as measured by the ADAS-Cog, an assessment scale sensitive to cognitive decline in AD (Kueper, Speechley and Montero-Odasso, 2018) where higher values indicate worse performance, (β = -0.282, 95% CI [-0.53, -0.04], p = 0.026; Figure 2F-G). With this, our replication analysis, directly reproduces population-based longitudinal results of Teipel et al. (2015). Critically, this protective effect substantially amplified specifically in AD. In the full ADNI cohort, the three-way interaction (age×genotype×amyloidosis) revealed that WT carriers with amyloid pathology experienced ∼50% slower AD-associated cognitive decline across the clinical spectrum (β = -0.31, l95% CI [-0.50, -0.11], p = 0.002; Figure 2H-I), demonstrating context-dependent cognitive resilience that becomes more evident under pathological conditions, providing indirect evidence of cognitive reserve (Stern *et al*., 2020).

### Regional specificity and systemic consequences of rs2765 variation

In order to understand the biological basis of these structural differences in the hippocampal and basal forebrain, we integrated structural AgeGain data with Allen Human Brain Atlas expression data. These analyses revealed regional mechanistic specificity underlying these structural effects. In hippocampal and basal forebrain regions, higher local *TACR3* expression predicted larger GMV (β = 0.10, p < 0.001) and rs2765 WT carriers showed significantly larger volumes (β = 0.10, p = 0.041; Figure 3A, left panel). This positive association was reversed in other brain regions, where *TACR3* expression negatively correlated with volume (β = -0.02, vs. β = -0.10 in hippocampus/basal forebrain regions, HippoBF, p < 0.001) and rs2765 genotype effects were absent (β = 0.04, p = 0.216; Figure 3A, right panel). These results were corroborated in the model estimated on all regional expression measurements, with the interaction between brain region and *TACR3* expression being highly significant (β = -0.20, p < 0.001; Supplementary Table 2), demonstrating that rs2765 structural effects occur mainly in regions where *TACR3* expression positively influences brain structure.

**Figure 3:**
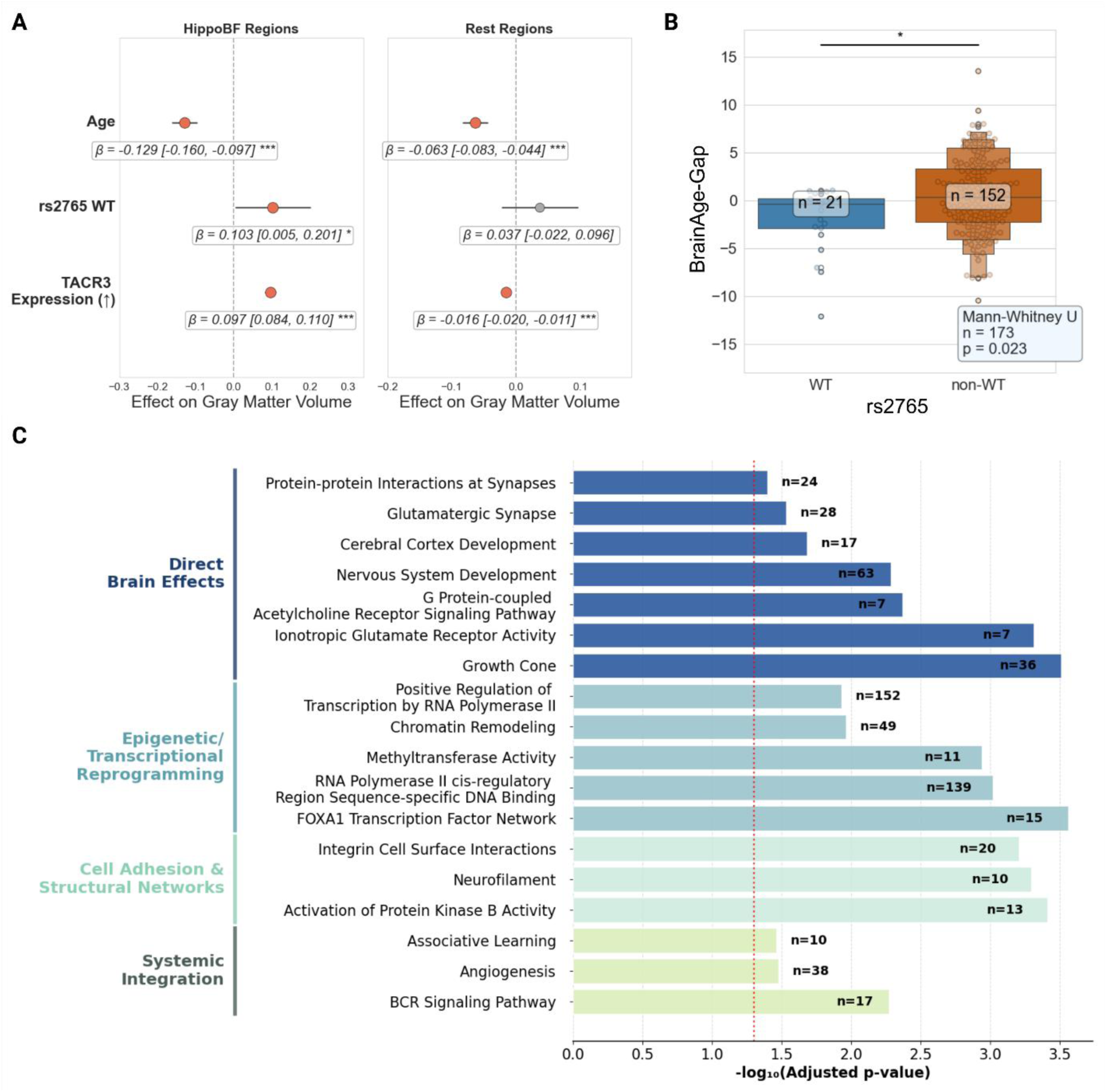
Convergent aging signatures associated with rs2765 genetic variation. **A)** Forest plots from region-specific analysis demonstrate functional specificity of rs2765 effects. In hippocampus/basal forebrain regions: *TACR3* expression positively predicts GMV (β = 0.10, p < 0.001) and rs2765 WT shows beneficial effects (β = 0.10, p = 0.0409). In contrast, other brain regions show a slightly negative *TACR3* expression-GMV relationship (β = -0.01, p < 0.001) and no rs2765 genotype effect (β = -0.037, ns), indicating that rs2765 structural benefits are specific to regions where *TACR3* expression positively correlates with brain volume specifically within the hippocampus and basal forebrain. **B)** Brain age gap (predicted brain age minus chronological age) showing significantly younger predicted brain age in rs2765 WT carriers (Mann-Whitney U test, p = 0.02, N = 173). **C)** Selected pathways from functional enrichment analysis of 228 pathways with >5 differentially methylated CpGs between rs2765 WT (n=18) and non-WT carriers (n=125), showing key pathways organized by mechanistic domain: Direct Brain Effects (dark blue), Epigenetic / Transcriptional Reprogramming (light blue), Cell Adhesion & Structural Networks (sage) and Systemic Integration (green). Bars represent -log₁₀(adjusted p-value); n = number of contributing CpGs.

To examine whether rs2765 effects extend beyond localized structural differences to influence global brain aging, we tested the hypothesis that WT carriers would show younger predicted brain age based on morphometry in the AgeGain cohort. Using a two-sided Mann-Whitney U test, WT carriers demonstrated significantly younger brain age compared to variant carriers (Hodges-Lehmann median difference: -1.96 years, 95% CI: [-3.89, -0.32], mean difference -2.3 years, 95% CI: [-3.99, -0.81], p = 0.023; effect size r = -0.31; Figure 3B). WT carriers showed brain age younger than chronological age (median: -0.38 years), while variant carriers showed brain age older than chronological age (median: +0.38 years, Supplementary Table 2).

To elucidate the molecular pathways that could coordinate both the regional structural benefits and systemic aging advantages, we performed genome-wide methylation analysis (based on blood samples) comparing WT and variant carriers in the AgeGain cohort. This analysis identified 2,313 differentially methylated CpG sites (FDR < 0.05) between WT and variant carriers, with functional enrichment revealing 228 significantly affected pathways (Supplementary Tables 3-4) with >5 contributing CpGs. The most statistically significant included FOXA1 transcription factor network, integrin cell surface interactions, and nervous system development.

Given the established role of NK3-R in neuronal excitability and aging, we selected 18 pathways spanning direct brain effects (nervous system development, growth cone), transcriptional reprogramming (RNA polymerase II regulation, FOXA1 network), structural networks (integrin interactions, neurofilament), and systemic integration (BCR signaling, angiogenesis) for detailed presentation (Figure 3C, see Methods for selection criteria). We next assessed whether these coordinated molecular changes translate to measurable differences in biological aging rates.

### rs2765 variant drives systemic aging acceleration through epigenetic mechanisms

To further investigate whether rs2765 effects extend beyond brain-specific changes to influence systemic aging processes, we examined associations based on the observed methylation patterns with PhenoAge, an epigenetic clock that captures accelerated biological aging and predicts mortality risk (Levine et al., 2018), in the AgeGain cohort. rs2765 WT carriers demonstrated markedly younger epigenetic age compared to variant carriers, with a clear gene-dose relationship across genotype groups (Figure 4A-B).

**Figure 4:**
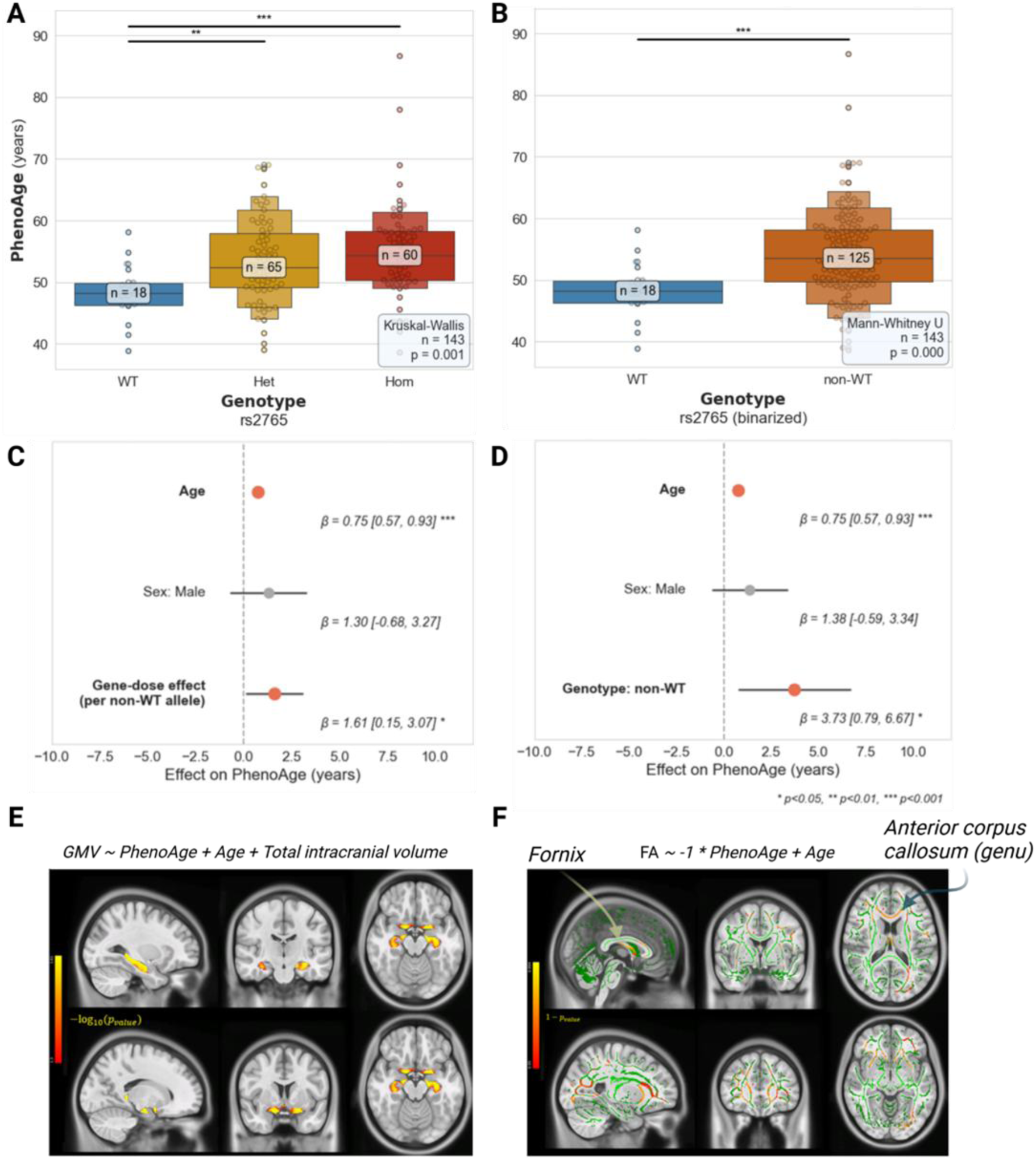
Associations between rs2765 genotype, PhenoAge, and brain structure. **A-B)** Distribution of PhenoAge values by rs2765 genotype in the AgeGain cohort. Panel **A** shows all three genotypes (WT, n=18; Het non-WT, n=65; Hom non-WT, n=60); Panel **B** shows binary classification (WT, n=18; non-WT, n=125). Box plots display median, quartiles, and individual data points. **C-D)** Forest plots from linear regression models predicting PhenoAge while controlling for chronological age and sex. Panel **C** shows gene-dose effect model (β per additional non-WT allele); Panel **D** shows binary genotype model (non-WT vs. WT reference). Error bars represent 95% confidence intervals. **E)** Masked voxel-wise analysis within hippocampal and basal forebrain brain regions of interest showing voxels, where higher GMV associates with lower PhenoAge (younger epigenetic age), controlling for chronological age, sex, and total intracranial volume. Statistical parametric maps show -log₁₀(p-values) from TFCE analysis, FDR-corrected at α = 0.05. MNI coordinates: upper row (-29, -18, -17), lower row (-17, 5, -17). **F)** White matter skeleton tract-based spatial statistics (TBSS) showing regions where higher fractional anisotropy associates with lower PhenoAge, controlling for age and sex. Green skeleton represents the white matter template; colored regions show FWE-corrected (1-p) values thresholded at α = 0.05. Prominent tracts include fornix, anterior corpus callosum (genu), and uncinate fasciculus. Statistical significance: *p < 0.05, **p < 0.01, ***p < 0.001.

Linear regression analyses controlling for chronological age and sex revealed robust associations in both gene-dose and binary models. Each additional non-WT allele was associated with 1.61 years of epigenetic age acceleration (95% CI [0.15, 3.07], p < 0.05; Figure 4C), while the binary comparison showed non-WT carriers exhibited 3.73 years older epigenetic age than WT carriers (95% CI [0.79, 6.67], p < 0.05; Figure 4D). These effect sizes are comparable to those observed for major lifestyle interventions and environmental exposures in epigenetic aging research (Fitzgerald *et al*., 2023; Ke, Lophatananon and Muir, 2024).

To validate PhenoAge as a meaningful aging biomarker in our neuroimaging context, we examined associations between peripherally assessed epigenetic age and brain structure in the AgeGain cohort. Voxel-wise analysis revealed that lower PhenoAge (younger epigenetic age) correlated with larger GMV in hippocampal and basal forebrain regions of interest that are also affected by rs2765 (Figure 4E). Similarly, tract-based spatial statistics demonstrated that lower PhenoAge associated with higher white matter integrity across multiple tracts, including the fornix and cingulum - critical pathways, through which the basal forebrain innervates to the hippocampus and cortical regions, as well as anterior corpus callosum and uncinate fasciculus (Figure 4F).

These converging findings establish that rs2765 variation influences systemic aging processes measurable through epigenetic mechanisms, with the brain structural correlates of PhenoAge supporting its validity as an aging biomarker that captures neurobiologically relevant age-related changes.

Having established that rs2765 influences both brain structure and systemic aging markers, we next investigated the molecular mechanism underlying these coordinated effects. The convergence of structural, cognitive, and epigenetic signatures suggests a fundamental biological pathway rather than isolated associations. Given rs2765’s location in the 3’UTR of the gene product, we hypothesized that post-transcriptional regulation underlies the variant’s pleiotropic effects.

### Post-transcriptional mechanisms explain rs2765 functional effects

The 3’UTR serves as a regulatory hub where multiple mechanisms such as miRNA binding, RNA-binding protein interactions, and secondary structure formation determine mRNA fate and protein expression. To systematically evaluate potential mechanisms, we analyzed the rs2765-containing region for G-quadruplex formation, miRNA binding sites, and RNA-binding protein motifs.

Analysis of conserved binding sites for miRNAs revealed only one miRNA predicted to bind the 3’UTR stretch of *TACR3*: hsa-miR-22-3p (Fig. 5A-B). However, the two predicted binding regions, in principle, did not cover the single nucleotide polymorphism (SNP) nor were they differing between WT and variant regarding the binding energy per se. Nevertheless, logistic probability (a proxy for the binding preference) was distinct and might hint at a different usage of both sides for miRNA hybridization. This might be also depending on slight structural effects that could be observed with a prediction algorithm (Fig. 5C). For G-quadruplex structures, no significant hints could be found (data not shown). However, analysis of potential RNA binding proteins’ motifs indicated that while the WT offers four positions for RBMX binding, the variant lacks the most proximal one, which is directly located at the SNP site (Fig. 5D). RBMX (an acronym of RNA Binding Motif protein, X-linked, also known as hn RNP G) is a 43 kDa X-chromosome encoded member of the heterogeneous nuclear ribonucleoprotein family (hn RNP), that is thought to act on RNA processing via splicing regulation but also other mechanisms (Elliott *et al*., 2019).

**Figure 5:**
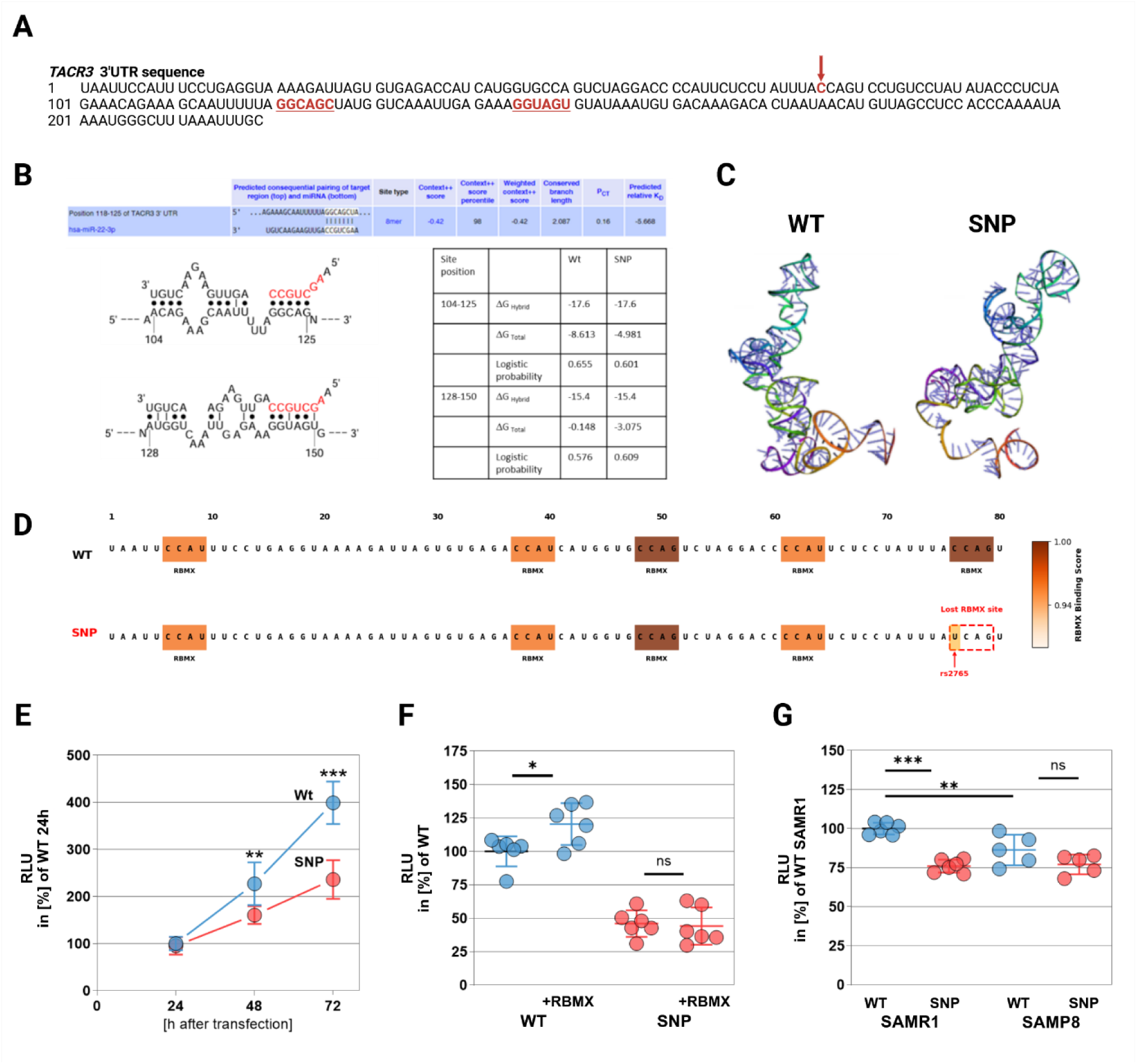
Mechanistic validation of rs2765-mediated post-transcriptional regulation of NK3-R protein expression. **A)** Sequence of *TACR3* 3’UTR with selected probable binding sites. The nucleotide affected by the polymorphism is labeled by a red arrow. **B)** Predicted, conserved miRNA binding to the *TACR3* 3’UTR. Two potential binding sites were identified in the human 3’UTR at position 104-125 and 128-150. Parameters for binding within the WT and the variant are indicated. **C)** Structure prediction for the two 3’UTR variants. **D)** RBMX binding sites with relative binding score within the two 3’UTR variants of *TACR3*. **E)** SIM-A9 murine microglia cells were transiently transfected with a reporter construct containing either the WT or the SNP variant. *Gaussia* luciferase activity from cell supernatant, reporting on translational activity, was collected for the indicated time points, analyzed and measured light emission normalized to protein content (relative light units, RLU). The experiment was conducted three times independently in triplicate (n=9). Statistical analysis was performed by using multiple Student’s t-test. **F)** Cells were transfected either with WT or variant reporter construct in combination with empty vector (-) or RBMX-encoding plasmid (+). Cell supernatant was collected after 72h and luciferase activity assessed as described. The experiment was conducted twice independently in triplicate (n=6). Statistical analysis was performed by using One Way ANOVA with Sidaks post-test. **G)** Brain slices from male mice (mean age 55 weeks) were taken into Transwell culture and transfected with the *TACR3* 3’UTR reporter constructs. 48h after transfection, hippocampal tissue was collected by punch biopsy and luciferase activity measured in the homogenate supernatant. Three animals were used with two slides per condition each (n=6). Statistical analysis was performed by using One-Way ANOVA with Sidaks post-test.

To further explore the functional consequences of the variant, we introduced a luciferase-based reporter construct in SIM-A9 cells as a mammalian cell system to validate RBMX-mediated post-transcriptional regulation. At 48 and 72 h post transfection, a significantly reduced enzyme activity could be measured in the cell supernatant, which indicates a reduced translational efficacy (Figure 5E) in the SNP (non-WT). Subsequently, co-transfection experiments with an RBMX expression vector revealed that only in the WT an elevation of reporter activity could be elicited by overexpression of the RNP while the SNP-carrying reporter did not react (Figure 5F). As the TACR3/NK3-R system has been associated with healthy aging in former studies and our own replicative human study, next, an analysis was performed on the basis of brain slice cultures from control mice (SAMR1, senescence accelerated aging mice resistant 1) and SAMP8 mice (senescence-accelerated mouse-prone 8). The latter is also thought to be applicable as a rodent sporadic Alzheimer disease model as it develops signs of neurodegenerative disease at higher age (Cheng, Zhou and Zhang, 2014; Liu, Liu and Shi, 2020). Interestingly, SAMP8 mice showed lower WT reporter activity in hippocampal area when compared to WT-reporter transfected SAMR1 slices, pointing at an already reduced translational activity which was also not responsive to RBMX activation (Figure 5F). The reporter activity between SAMP8 WT and SNP variant was insignificant and comparable to the reduced activity found in SAMR1 slices transfected with the SNP-containing reporter (Figure 5G).

### Cross-species validation confirms TACR3-mediated aging effects

As the SAMP8-derived brain slices already pointed at a reduced gene expression efficacy of the *TACR3* gene, the endogenous protein expression of NK3-R and RBMX were analyzed (Figure 6A-C): about 30% less NK3-R protein could be detected in the hippocampus of SAMP8 mice when compared to SAMR1 controls, while RBMX was indifferent. Additionally, the amount of RBMX and NK3-R protein showed a positive correlation (Figure 6D). Finally, relative hippocampal weight positively correlated with NK3-R protein amount while this could not be observed for hypothalamus (Figure 6E, data for hypothalamus not shown). Together with the negative correlation between NK3-R protein levels and acetylcholine esterase activity (AChE; Figure 6F), this points to functional consequences in the rodent model of accelerated aging and highlights the importance of the TACR3/NK3-R system for cholinergic signaling in the aged brain.

**Figure 6:**
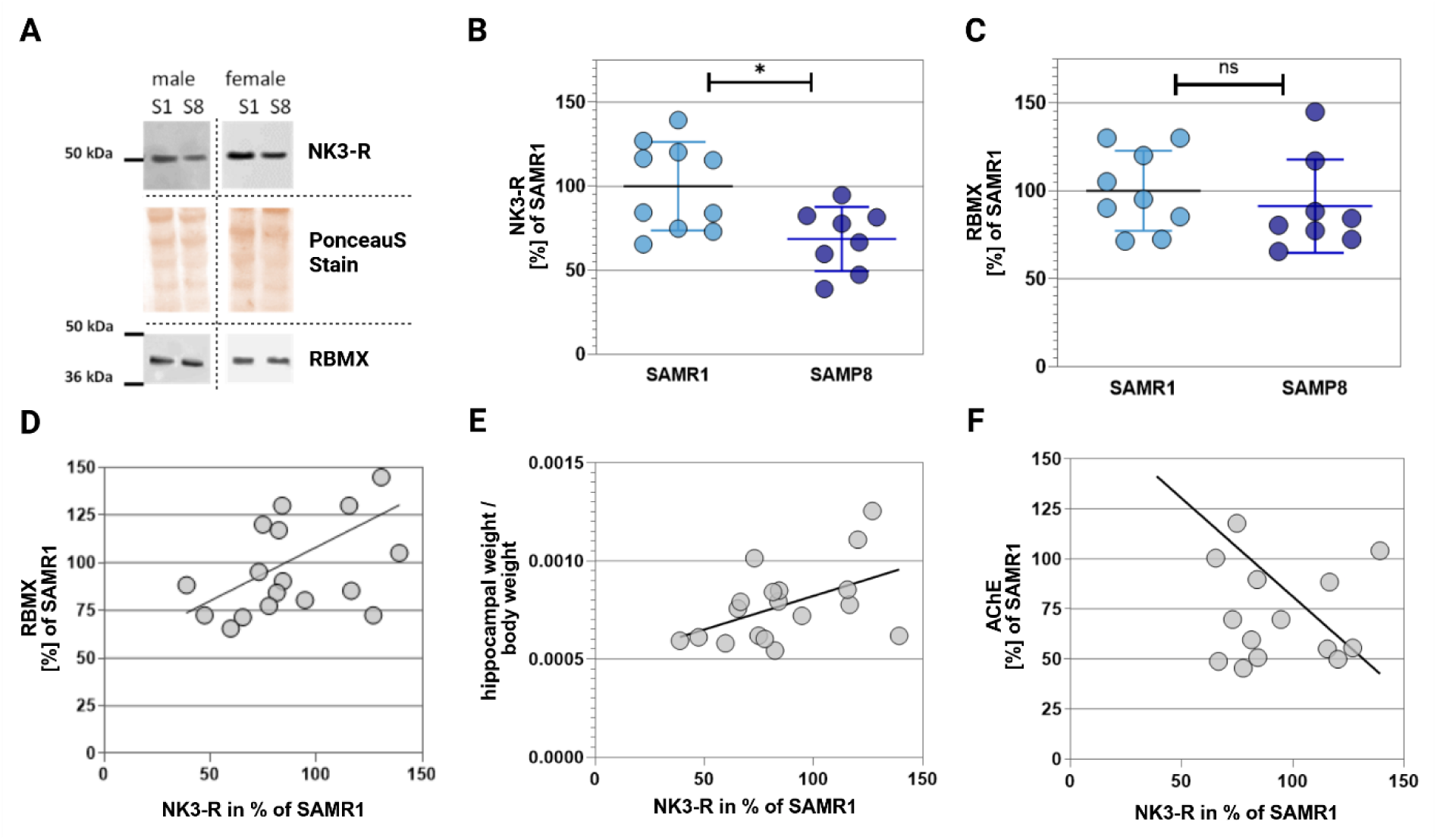
NK3-R expression associates with hippocampal mass and cholinergic function. **A)** Western blots were performed to assess the amount of NK3-R and RBMX in the hippocampus of SAMR1 and SAMP8 mice. Forty µg of protein were subjected to 10% SDS PAGE and transferred to nitrocellulose membrane. Protein-dependent signals were measured by HRP-labelled secondary antibodies and a CCD camera. Exemplary images are shown for male and female mice of both strains. In total 10 SAMR1 mice were used (5 male, 5 female) and 9 SAMP8 mice (4 male and 5 female). All mice were aged 35 to 42 weeks. Ponceau S red stain served as a loading control. **B)** Quantitation of NK3-R from the density signals obtained as shown in A. **C)** Quantitation of RBMX. For B and C unpaired Student’s t-test was used. **D)** Correlation analysis of RBMX and NK3-R in all mice tested (n=19). Pearson r= 0. 4581, one-sided p = 0.0280. **E)** Correlation analysis of relative hippocampal weight and NK3-R protein amount. Pearson r= 0.4882, one-sided p = 0.0199. **F)** Correlation analysis of AChE activity and NK3-R protein amount. Pearson r= -0.5515, one-sided p = 0.0088.

## Discussion

The current findings provide a mechanistic characterization of rs2765 as a 3’UTR variant conferring aging resilience in humans. Unlike previous aging-associated variants identified through genome-wide approaches, rs2765 demonstrates a complete causal pathway from genetic variation through post-transcriptional regulation to systemic aging outcomes. The convergence of brain structural, cognitive, and epigenetic aging signatures, combined with prior therapeutic validation demonstrating that NK3-R agonists enhance memory in aged animals (de Souza Silva *et al*., 2013), positions *TACR3* and its rs2765 variant as both a precision aging biomarker and an immediately actionable therapeutic target for promoting healthy aging.

The systematic validation, further characterization in human cohorts and follow-up translational experiments suggest the *TACR3* variant rs2765 as a mechanistically characterized determinant of aging resilience. Luciferase assays and cross-species validation demonstrate that this 3’UTR variant ameliorates RBMX-mediated post-transcriptional regulation, reducing NK3-R protein expression and providing a molecular mechanism for the brain structural, cognitive, and systemic aging associations replicated across independent human cohorts (AgeGain and ADNI) and complementary aging mouse models (SAMR1/SAMP8). Unlike most aging-associated variants identified through genome-wide approaches, which remain mechanistically uncharacterized and lack replication, rs2765 combines experimental mechanistic validation with cross-cohort and translational evidence, establishing a framework for mechanism-based precision aging medicine.

### Post-transcriptional control of aging: From 3’UTR variants to systemic effects

The location of rs2765 in *TACR3*’s 3’UTR provides important mechanistic insights, as 3’UTR variants are up to 16-fold more likely than other non-coding variants to have phenotypic effects through post-transcriptional regulation (Romo, Findlay and Burge, 2024). Supporting this potential, a systematic genome-wide functional screen of 12,173 3’UTR variants using massively parallel reporter assays identified rs2765 among variants with significant post-transcriptional effects, demonstrating activity specifically in lymphoblastoid cells (GM12878; log2FC = -0.74, p < 0.05) across six human cell lines tested (Griesemer *et al*., 2021). Notably, this B-cell-specific functional effect aligns with our genome-wide methylation analysis, where B-cell receptor signaling pathways were significantly enriched among differentially methylated regions in rs2765 carriers. This convergence suggests that rs2765’s systemic aging effects may involve immune system modulation beyond its brain-specific actions.

This mechanism is particularly relevant to aging biology, as RNA-binding proteins that recognize 3’UTR motifs become increasingly dysregulated with senescence and contribute to transcriptome-wide splicing alterations in cellular aging (Dong *et al*., 2018; Varesi *et al*., 2023). Our identification of RBMX binding site disruption by rs2765 exemplifies this pathway: RBMX functions as a master regulator maintaining chromatin state and controlling expression of chromatin modifier *CBX5* in myeloid cells (Prieto *et al*., 2021).

The 3.7-year epigenetic age difference (95% CI [0.79, 6.67]) between WT and non-WT represents a substantial genetic effect on biological aging. For context, the habit of smoking shows 2.1-2.3 years PhenoAge acceleration in older adults (Cardenas *et al*., 2022), while combined in utero tobacco exposure plus childhood smoking initiation results in 2.89 years (95% CI [2.75, 3.03]) (Cui *et al*., 2024). Lifestyle interventions report 2.04-3.23 years reduction on the Horvath clock (Fitzgerald *et al*., 2023) and 1.16 years PhenoAge reduction with regular physical activity (Wu *et al*., 2024). Notably, PhenoAge demonstrates weaker associations with environmental factors compared to GrimAge, making this genetic effect particularly noteworthy (Cardenas *et al*., 2022; Cordero *et al*., 2024). However, given the limited sample size in AgeGain (n=18 WT carriers), replication in larger independent cohorts is warranted. This systemic aging signature is paralleled by younger predicted brain age in WT carriers (-2.3 years, 95% CI [-3.99, -0.81], p=0.023), suggesting coordinated effects spanning systemic to neural aging processes. Notably, rs2765 alone explains 2.8% of PhenoAge variance in our cognitively healthy cohort, exceeding the combined contribution of 11 top SNPs (1.86%) identified in large-scale epigenetic clock GWAS studies (Morales Berstein *et al*., 2022). This substantial single-variant effect, validated from molecular mechanism through systemic aging outcomes, positions rs2765 as among a promising genetic determinants of biological aging.

The substantial methylation reprogramming (2,313 CpGs, 228 pathways) likely reflects TACR3/NK3-R’s pleiotropic roles across multiple systems. NK3-R is expressed not only in hippocampal and cholinergic neurons but also in immune cells, vascular endothelium, and peripheral tissues, coordinately influencing transcriptional programs governing inflammation, angiogenesis, and metabolic regulation (Zhang, Wang and Chu, 2020). Pathway enrichments directly relevant to neuronal function and acetylcholine signaling support aging-specific mechanisms beyond potential reproductive effects.

### Regional specificity and the “excited or not” hypothesis

The regional specificity of rs2765 effects provides crucial mechanistic insight into NK3-R function across brain circuits. Our Allen Brain Atlas validation demonstrates that rs2765 GMV benefits occur primarily in regions where *TACR3* expression positively correlates with brain volume, namely in the hippocampal and basal forebrain areas, while regions showing negative *TACR3*-volume correlations show no rs2765 genotype effects. This supports the “excited or not” hypothesis, where NK3-R serves as a neuronal excitability control unit with fundamentally different roles across brain circuits (Zhang, Wang and Chu, 2020).

The mechanistic basis for this regional specificity involves NK3-R’s control of neuronal excitability through regulation of ion channels including inhibition of G protein-coupled inwardly rectifying potassium (GIRK) channels and activation of transient receptor potential cation 4/5(TRPC4/5) channels (Boyle *et al*., 2022). In the hippocampus specifically, *TACR3* directly regulates synaptic plasticity by controlling spine density, pruning, and long-term potentiation through Ca^2+^/calmodulin-dependent protein kinase II (CaMKII) signaling (Wojtas *et al*., 2024) and is implied in the hippocampal neurogenesis in the context of AD (Koca *et al*., 2024). The validation in SAMR1/SAMP8 mice is consistent with this mechanism, showing 30% reduced NK3-R protein in accelerated aging mice hippocampus. This reduction correlated with both, decreased hippocampal weight (r=0.49) and acetylcholine esterase activity (r=-0.55), linking NK3 receptor expression to structural and functional outcomes.

Additionally, hippocampal activation patterns and effective functioning of memory circuits depend on precisely timed acetylcholine secretion from cholinergic basal forebrain neurons, which innervate the hippocampus via the fornix (Boskovic *et al*., 2019). Our findings that fornix structural connectivity is better preserved in biologically younger individuals (Figure 4F), combined with WT carriers being on average 3.7 years younger as measured by PhenoAge, suggest that more sustained or effectively timed neurotransmitter release may be an additional mechanism explaining functional and structural outcomes.

Given the functional outcome findings for the clinical spectrum of AD, where WT-presence moderates the impact of amyloidosis on cognition, these neurobiological mechanisms support an active resilience framework. Human WT carriers demonstrate preserved cognitive function, particularly during pathology accumulation, showing ∼50% slower decline in amyloid-positive individuals (Figure 2H-I). These findings align with conceptions of resilience being an active, independent biological process rather than a passive capacity or counteracting the pathological process (e.g. amyloid proteostasis) itself (Stern *et al*., 2020; Nestler and Russo, 2024)..

Hence, rs2765 represents a mechanistically validated resilience variant: WT carriers do not prevent amyloid accumulation (no brain maintenance effect; genotype frequencies similar across amyloid-positive and amyloid-negative subgroups, Supplementary Table 1) but rather maintain cognitive capacity through active compensatory mechanisms that become most evident under pathological stress. This mechanistic dissociation between preservation of function without prevention of pathology, mirrors the adaptive mechanisms observed in stress resilience (Nestler and Russo, 2024), where protective responses are actively induced rather than avoiding damage. The resilience conferred by rs2765 operates through multiple biological substrates: larger hippocampal and basal forebrain volumes provide structural buffering (brain reserve), while enhanced cholinergic modulation, increased synaptic plasticity, and potentially preserved hippocampal neurogenesis (Koca *et al*., 2024) enable active functional compensation when pathology accumulates (cognitive reserve). Critically, the protective effect amplifies specifically when pathology burden increases, which constitutes a hallmark of cognitive reserve mechanisms where compensation becomes most relevant under stress. Hippocampal volume as a factor of cognitive reserve has been demonstrated previously by our group (Wolf, Fischer, Fellgiebel, and for the Alzheimer’s Disease Neuroimaging Initiative, 2019), and hippocampal hypertrophy in early stages of dementia has been discussed as a compensatory mechanism (Chételat *et al*., 2010; Le Stanc *et al*., 2024), providing converging evidence that the structural advantages observed in WT carriers support functional compensation under pathological stress.

This resilience framework is further supported by recent demonstrations that cognitively healthy centenarians are enriched for protective alleles across multiple AD-associated loci, with effect sizes amplified 1.78-fold relative to case-control studies (Tesi *et al*., 2024), suggesting polygenic resilience mechanisms operate at extreme longevity. rs2765 represents a rare example of such resilience variant with complete mechanistic characterization from molecular pathway (RBMX-dependent post-transcriptional regulation) through cellular validation (luciferase assays, brain slice cultures) to systemic aging outcomes (epigenetic age, brain age, coordinated methylation reprogramming).

Unlike *APOE* ε4, which modulates both AD risk and progression rates (Cosentino *et al*., 2008; Whitehair *et al*., 2010; Suzuki *et al*., 2020), rs2765 primarily influences resilience to age-related and pathology-associated decline without clear effects on amyloid pathology itself. This mechanistic distinction has important implications for precision medicine: APOE genotyping stratifies amyloid accumulation risk, while rs2765 genotyping predicts functional consequences of accumulated pathology. While comparing an endogenous genetic protective mechanism operating across the lifespan with time-limited pharmacological interventions requires appropriate caution, the ∼50% slowing of cognitive decline observed in amyloid-positive WT carriers is of similar or greater magnitude than clinical benefits demonstrated with recently approved anti-amyloid monoclonal antibodies, lecanemab (26% slowing on ADAS-Cog; Dyck *et al*., 2023) and donanemab (19-35% slowing on ADAS-Cog depending on tau stratification; Sims *et al*., 2023). This similarity in effect magnitude suggests that NK3-R pathway modulation, whether through preserved endogenous signaling in WT carriers or potential therapeutic agonists, may represent a physiologically significant target for cognitive resilience. With anti-amyloid therapies now approved and increasing the frequency of amyloid screening, rs2765 genotyping could complement APOE-based risk assessment by identifying individuals who, despite developing amyloidosis, are most likely to maintain cognitive function (WT carriers) versus those at highest risk for rapid functional decline (variant carriers). This distinction becomes particularly relevant as the field shifts from preventing amyloid accumulation to managing its functional consequences.

### Clinical implications: NK3-R as a bidirectional therapeutic target

Our mechanistic validation of the NK3-R pathway establishes it as a promising therapeutic target with immediate clinical validation opportunities. Multiple independent studies demonstrate that NK3-R agonism rescues memory decline in rodent AD models via cholinergic mechanisms (de Souza Silva *et al*., 2013; Koca *et al*., 2023, 2024; Huang *et al*., 2024). This therapeutic potential, combined with our demonstration that rs2765 genotype determines differential aging trajectories (1.61 years epigenetic acceleration per variant allele), positions genotype-stratified NK3-R modulation as a precision medicine approach for cognitive aging interventions.

The mechanistic implications for NK3-R pharmacology are noteworthy. Given that WT carriers show cognitive resilience and higher NK3-R expression, while variant carriers show accelerated aging and reduced NK3-R, the directionality suggests that NK3-R agonism would be protective - as demonstrated by senktide in preclinical models (de Souza Silva *et al*., 2013; Huang *et al*., 2024).

In turn, NK3-R antagonism might phenocopy the variant allele’s effects, raising questions about long-term cognitive safety of fezolinetant, an FDA-approved NK3-R antagonist for postmenopausal vasomotor symptoms. While multiple phase 3 trials demonstrate fezolinetant’s efficacy and acceptable short-term safety profile over 12-52 weeks (Johnson *et al*., 2023; Lederman *et al*., 2023; Neal-Perry *et al*., 2023; Menegaz de Almeida *et al*., 2025), none assessed cognitive function as an endpoint. The temporal separation between typical treatment initiation (mean age ∼50-55) and dementia onset (>65-70) means that potential long-term cognitive effects remain unexamined in current clinical trials.

Genotype-stratified monitoring of fezolinetant users could determine whether rs2765 modifies long-term cognitive trajectories following NK3-R antagonism, while also validating whether genetic background predicts differential responses to NK3-R modulation. Similarly, rs2765 genotyping may inform clinical trial enrollment for NK3-R agonist interventions, enabling targeted patient selection for individuals most likely to benefit from enhanced cholinergic signaling.

The convergence of mechanistic validation, cross-cohort replication, and existing pharmacological tools positions rs2765 as an immediately actionable biomarker. Unlike most aging-associated genetic variants that lack therapeutic translation, the availability of both FDA-approved NK3-R antagonists and preclinical agonist validation provides a unique opportunity for rapid clinical implementation in precision aging medicine.

### Limitations and Implementation Challenges

Cross-sectional cognitive assessment in AgeGain detected no significant rs2765 association, likely reflecting limited power (n=25 WT carriers) and the inherent limitations of cross-sectional designs for detecting gradual cognitive decline trajectories. Significant effects emerged only in longitudinal ADNI analyses (Figure 2F-I), consistent with a resilience mechanism detectable through decline rates rather than single-timepoint comparisons.

While AgeGain’s sample size (n=188; 25 WT carriers) limits power for genetic associations, ADNI validation (n=809; 111 WT carriers) and our subsequent mechanistic studies strengthen causality arguments. The pathology-dependent effect amplification observed in ADNI (∼50% cognitive protection in amyloid-positive WT carriers) therefore requires replication in independent cohorts with comprehensive biomarker characterization.

The European ancestry focus requires validation across diverse populations. Given only about 12.8% of Europeans are WT carriers (*dbSNP rs2765*, 2025), potential evolutionary trade-offs may exist, possibly related to NK3-R’s role in reproduction (Zhang, Wang and Chu, 2020), though this remains speculative. Although linkage disequilibrium could theoretically implicate nearby variants, de Souza Silva et al. (2013) explicitly identified rs2765 as the functional variant through targeted sequencing, and our luciferase experiments directly tested the rs2765 variant itself, supporting rs2765 as the causal variant.

Our mechanistic validation has methodological constraints. While luciferase reporter assays demonstrate RBMX-mediated regulation of the rs2765 variant, these experiments used microglia (SIM-A9) rather than neurons. Although *TACR3* is expressed in both cell types, neuronal validation would strengthen mechanistic specificity. The SAMP8 mouse model demonstrates NK3-R’s role in aging but cannot directly model the human rs2765 variant. Blood-based methylation analyses serve as systemic proxies but may not fully reflect brain-specific processes.

Multiple testing within neuroimaging and methylation outcomes was addressed through FDR correction, though further independent replication remains essential. While effect sizes are statistically robust, their individual-level clinical significance requires prospective validation in intervention studies.

## Conclusion

We establish rs2765 as a mechanistically characterized determinant of aging and AD resilience, demonstrating an active resilience mechanism from RBMX-dependent post-transcriptional dysregulation to brain structure, cognition and systemic aging. The convergence of structural, cognitive, and epigenetic evidence, including ∼50% cognitive protection in amyloid-positive WT carriers, positions rs2765 and the TACR3/NK3-R system as both biomarker and therapeutic target. With NK3-R antagonists now in clinical use, understanding how rs2765 genotype influences treatment outcome becomes increasingly urgent. This work exemplifies how systematic mechanistic validation can bridge genetic associations into actionable clinical insights, providing a framework for precision geromedicine.

## Methods

### 1. Study design and participants

#### 1.1 AgeGain cohort

The present study utilized data from the AgeGain study, a multicentric, multimodal imaging, interventional, longitudinal, parallel group. The study was preregistered with the German Clinical Trials Register (DRKS, ID: DRKS00013077) on November 19, 2017 (Wolf *et al*., 2018).

A total of 235 cognitively healthy elderly subjects aged over 59 years were recruited at three sites: the University medical centers in Mainz and Rostock, and the German Sport University Cologne between 2016 and 2019. Recruitment was conducted through newspaper announcements and flyers.

Written informed consent was obtained from all participants, and the study protocol was approved by the respective local ethics committees. Exclusion criteria included psychological, neurological, or cognitive illnesses; cardiovascular disease; disorders restricting physical capacity; diabetes; medication affecting cognitive performance; insufficient German language skills; concurrent participation in other trials; and MRI contraindications. Further details about the study design have been published previously (Wolf *et al*., 2018).

##### Study sample and data availability

For the present study, we analyzed multiple data modalities from the AgeGain cohort. From the total cohort, 188 participants had available rs2765 genotype information. Of these 188 genotyped participants, all had VLMT7 cognitive outcome assessments, 173 had MRI data and 143 had MRI and methylation data. A more detailed overview on sample size and cohort description can be found in Supplementary Table 1.

#### 1.2 ADNI cohort

Data used in the preparation of this article were obtained from the Alzheimer’s Disease Neuroimaging Initiative (ADNI) database (adni.loni.usc.edu). The ADNI was launched in 2003 as a public-private partnership, led by principal investigator Michael W. Weiner, MD. The original goal of ADNI was to test whether serial MRI, positron emission tomography (PET), other biological markers, and clinical and neuropsychological assessment can be combined to measure the progression of mild cognitive impairment (MCI) and early Alzheimer’s disease (AD). The current goals include validating biomarkers for clinical trials, improving the generalizability of ADNI data by increasing diversity in the participant cohort, and to provide data concerning the diagnosis and progression of Alzheimer’s disease to the scientific community. For up-to-date information, see adni.loni.usc.edu. Written informed consent was obtained from all participants, and the study protocol was approved by the respective local ethics committees.

##### Study Sample and Data Availability

For the present analysis, we included 809 participants aged 55 to 92 years with available rs2765 genotyping. Of these, 284 were characterized as cognitively normal at baseline, 480 as patients of mild cognitive impairment (MCI) and 48 as patients with dementia based on the diagnostic inclusion criteria of the ADNI (https://adni.loni.usc.edu/wp-content/uploads/2008/07/adni2-procedures-manual.pdf). For a cognitive outcome subanalysis, we defined a subgroup of these subjects as healthy aging, selecting cognitively normal subjects with negative amyloid PET throughout enrollment yielding 121 subjects.

Of the 809 genotyped subjects, 799 had data of automatically segmented hippocampal and intracranial volume available, whereas 729 had data of amyloid-PET assessments and cognitive performance available. A more detailed overview on sample size and cohort description can be found in Supplementary Table 1.

### 2. Genotyping

For the AgeGain cohort, DNA extraction and genotyping procedures were performed as previously described (Wolf et al., 2018). Briefly, rs2765 genotyping in the *TACR3* gene was performed using PCR followed by pyrosequencing on a PyroMark system. Participants were classified based on rs2765 genotype as WT (GG), heterozygous (AG), or homozygous variant (AA), consistent with previous studies (de Souza Silva *et al*., 2013; Teipel *et al*., 2015) and the 1000 Genomes Project and dbnSNP classifications, respectively (1000 Genomes Project Consortium *et al*., 2015; *dbSNP rs2765*, 2025).

For the ADNI cohort, details of genome sequencing procedures can be accessed online (https://adni.loni.usc.edu/wp-content/uploads/2010/09/ADNI_WGS_Notice_20130917.pdf).

### 3. Data acquisition and processing

#### 3.1 Neuroimaging assessments

##### 3.1.1 MRI data acquisition - AgeGain

For the AgeGain cohort, identical T1- and diffusion-weighted structural MRI sequences were implemented on three Siemens 3T Scanners at the three study sites: on a Prisma in Cologne, a Trio in Mainz and a Verio in Rostock. T1-weighted images were acquired using a Generalized Autocalibrating Partial Parallel Acquisition sequence at 1900ms TR, 2.52ms TE and isotropic voxel size of 1mm^3. Diffusion weighted imaging (DWI) was acquired using an echo planar multiband sequence, multiband factor 3, TR 5500ms, TE 104ms and isotropic voxel size of 2mm^3. 64 diffusion gradients were acquired for b=1000 and for b=2000, as well as two b=0 images. To estimate distortion correction, we additionally acquired one b=0 image, where phase encoding had been inverted.

##### 3.1.2 MRI data acquisition - ADNI

For the ADNI cohort, several T1-weighted structural MRI sequences were employed at different scanners. To improve comparability of the data for voxelwise analyses, we included only the first scans for each subject that had completed the preprocessing and normalization pipeline (https://adni.loni.usc.edu/data-samples/adni-data/neuroimaging/mri/mri-pre-processing/).

##### 3.1.3 PET acquisition and analysis

PET analyses of the ADNI cohort were conducted at various sites using various scanners and subjects were administered either Pittsburgh Compound B (PIB) or florbetapir (AV45) tracers. Subsequently, scans were extensively processed with the goal of normalization and inter-site comparability by the ADNI PET core facility (https://adni.loni.usc.edu/data-samples/adni-data/neuroimaging/pet/). These efforts were enhanced by the Alzheimer’s Disease Sequencing Project – Phenotype Harmonization Consortium (https://adsp.niagads.org/funded-programs/phenotype-harmonization/), who provide a dichotomous classification of amyloid PET scans into cerebral amyloidosis negative or positive to the public. Relying on this scan-wise classification, we dichotomized the ADNI sample for cognitive outcome analyses in the following way: subjects were either added to the group of present or developing amyloidosis if any of the assessed amyloid PET scans was classified as amyloid positive, or into the group of no present or developing amyloidosis otherwise.

##### 3.1.4 Image preprocessing

###### Structural MRI analysis

For voxelwise GMV analyses both in the AgeGain and ADNI cohorts, T1-weighted images were spatially normalized and tissue segmented and total intracranial volume (TIV) calculated using the SPM12 toolbox CAT12 (Ashburner *et al*., 2014; Gaser *et al*., 2024). Voxel-wise statistical analyses were calculated using the Threshold Free Cluster Enhancement toolbox for SPM12 (https://neuro-jena.github.io/software.html#tfce). Hippocampus and basal forebrain masks were derived from the neuromorphometrics atlas included in CAT12.

For the ADNI cohort, the hippocampal and intracranial volumes were calculated at ADNI core facilities using automatic image segmentation implemented in FreeSurfer (Hartig *et al*., 2014).

###### Diffusion MRI analysis

For diffusion MRI data in the AgeGain cohort, Eddy from FSL 6.0.4 was used to correct spatial distortions, eddy current artefacts and movement (Andersson and Sotiropoulos, 2016). Diffusion tensors were fitted to the data using MRTRIX 3.0.2. Subsequently, fractional anisotropy (FA) values were calculated from the tensors’ eigenvalues. The individual FA maps were spatially normalized using TBSS from FSL. Voxelwise statistics were calculated within TBSS using threshold free cluster enhancement (TFCE).

###### Brain age estimation5

To estimate subjects’ age based on structural brain scans, we employed the BrainAge toolbox (Kalc *et al*., 2024). The Gaussian process regression based estimator was trained using brain scans of the training sample provided by Kalc et al. (https://github.com/ChristianGaser/BrainAGE) with a minimum age of 40 years. As we were interested in associations of “younger” or “older” brains with rs2765, we calculated the residual estimation error “BrainAge Gap” as the difference between the estimated and the actual age.

#### 3.2 Cognitive assessment

In AgeGain, verbal learning and memory were assessed using the Verbal Learning and Memory Test (VLMT), with delayed recall (trial 7) as the primary outcome measure. In ADNI, cognitive function was assessed using the 13-item Alzheimer’s Disease Assessment Scale-Cognitive subscale (ADAS-Cog13), where higher scores indicate worse performance. Assessments were conducted longitudinally according to the ADNI protocol (Mohs, Rosen and Davis, 1983).

#### 3.3 DNA methylation

DNA methylation was assessed in 143 samples from the AgeGain cohort. For methylation profiling, genomic DNA samples were processed using the Illumina Infinium MethylationEPIC BeadChip array (v1.0 B4, ∼850,000 CpG sites) according to the manufacturer’s protocol.

##### 3.3.1 Pre-processing pipeline

Raw methylation data were processed using ChAMP (Tian *et al*., 2017) version 2.28.0 in R (version 4.2.2) with a two-branch strategy: (1) For epigenetic clock analyses, beta values were extracted immediately after import to preserve maximum CpG coverage required for clock algorithms; (2) For differential methylation analyses, comprehensive quality control included (i) removing probes with detection p-value >0.01 in more than 10% of samples, (ii) removing probes with insufficient bead count in > 5% of samples, (iii) removing cross-reactive, SNP-containing and sex chromosome probes, (iv) BMIQ normalization for probe-type bias correction, and (v) ComBat batch correction adjusting for sex, age, slide, and array position (Zhang, Parmigiani and Johnson, 2020). After quality control, 717,800 CpG sites across 143 samples were retained for differential methylation analysis.

#### 3.4 Gene expression data (Allen human brain atlas)

*TACR3* expression data were extracted from the Allen Human Brain Atlas (Hawrylycz *et al*., 2012), which provides genome-wide transcriptional profiles across 3,702 samples from six neurotypical adult brains. Expression data were normalized using robust multi-array average (RMA) preprocessing and log₂-transformed. For regions with multiple samples, expression values were averaged within each donor brain.

### 4. Analysis of structure-function relations of rs2765

#### 4.1 Structural and functional analyses of *TACR3* 3’UTR (*in silico*)

A 178-nucleotide sequence encompassing rs2765 was analyzed, corresponding to positions 1706-1878 of the *TACR3* transcript (NM_001059.3).The rs2765 variant (chr4:103589609, GRCh38) corresponds to position 76 in the analyzed sequence, where the reference allele contains cytosine (C) and the variant allele contains uracil (U) in the RNA sequence.

The QUFIND module (http://www.soodlab.com/qufinder/) was used to identify probable G-quadruplexes in the selected RNA stretch. TargetScan (https://www.targetscan.org/vert_80/) was applied to assess potentially binding miRNAs and STarMir (https://sfold.wadsworth.org/cgi-bin/index.pl) to evaluate binding probability. trRosettaRNA was used for 3D structure prediction (Wang *et al*., 2023).

#### 4.2 RBMX binding prediction (*in silico*)

RNA-binding protein motif analysis was performed to predict the functional impact of the rs2765 G>A variant on post-transcriptional regulation.

RNA-binding protein motif scanning was performed using the RNA-Binding Protein DataBase (RBPDB) to identify predicted binding sites across both sequence variants (Cook *et al*., 2011). RBMX binding sites were predicted using position weight matrices with binding probability scores ranging from 0.80 to 1.00, with higher scores indicating stronger predicted binding affinity.

The analysis revealed that the rs2765 variant disrupts a high-probability RBMX binding site (score = 1.00) present in the reference sequence, providing the rationale for subsequent experimental validation of RBMX-mediated regulation.

#### 4.3 3’UTR reporter plasmid construction

Based on a WT and a rs2765 variant-containing DNA derived from blood samples of the AgeGain study population, the respective 3’UTR of the TACR3 gene was amplified by using the FailSafe Kit (buffer E; Biosearch Technologies) and the TACR3_3’-UTR_rev und TACR3_3’-UTR oligonucleotides (5‘ GCGGCCGCTAATTCCATTTCCTGAGGTA AAAG 3‘; 5‘ GCGGCCGCAAATTTAAAGCCCATTTTATTT TGG 3‘). The obtained amplicons were firstly integrated in a modified pUC19 vector (Clontech, Mountain View, CA., USA, modification of polylinker by R. Postina, JGU Mainz), from where they were excised by the flanking *Not*I recognition site and finally inserted in the pCMVGLuc plasmid (NEB Biolabs). Identity of the inserted genomic sequences was proven by *Bsr*I restriction (lack of one cleavage site in the rs2765 variant) and by sequencing (Eurofins Scientific SE (Luxemburg, LU)). All plasmids were produced in DH5a *E. coli* (NEB Biolabs) and purified in endotoxin-free quality (Endo free Plasmid Midi Kit, Qiagen).

#### 4.4 Cell culture experiments

SIM-A9 cells (Nagamoto-Combs et al., 2014, Applied Biological Materials Inc.; Richmond (BC), Canada) were maintained at 37 °C, 95% humidity, and 5% CO2 using Dulbecco’s Modified Eagle Medium/Nutrient Mixture F-12 (DMEM:F-12), supplemented with 10% v/v heat-inactivated fetal bovine serum (Gibco, Carlsbad, CA, USA), 5% v/v heat-inactivated donor horse serum (Gibco, Carlsbad, CA, USA), 1% v/v Penicillin-Streptomycin (Sigma-Aldrich; Steinheim, Germany), and 1% v/v L-Glutamine (Sigma-Aldrich; Steinheim, Germany). Upon reaching 80% confluency (two times a week), the cells were passaged by adding Trypsin/EDTA-solution (Sigma-Aldrich; Steinheim, Germany) for 5 minutes at 37 °C, 95% humidity, and 5% CO2.

SIM-A9 cells were seeded at 20.000 cells per well on transparent 96 well plate (Greiner). After 24h of cultivation, cell supernatant was aspirated and 100 µl serum-free medium added per well. FuGENE® HD Transfection Reagent (Promega)-DNA-Mix was prepared by using 100 ng of DNA (0.1 µl) per well and 0.4 µl of reagent in 4.5 µl serum-free medium. The mixture was incubated for 15 min at RT with an initial shaking of 30 s and then added to the cells. After 5h incubation at 95% humidity and 5% CO2, 37°C, the serum-free medium was exchanged with 100 µl culture medium.

At the indicated time points, 10 µl of cell supernatant (containing the secreted *Gaussia* luciferase as a proxy for translational activity) were aspirated, pipetted on white 96 well plates (Greiner) and stored at -20°C until measuring. Per well, 40 µl of Renilla luciferase assay kit (Promega) were added and luminescence detected by a fluorimeter (FLUOStar Omega). For normalization of signals, the remaining cell supernatant was aspirated from the culture plates, the cells lysed in passive lysis buffer (Promega) and incubated for at least 15min at -20°C to support cell lysis. Protein content was measured by a Bradford assay with the Roti Nano quant reagent (Roth).

For co-transfection experiments, the hnRNP G (RBMX) (NM_002139) Human Tagged ORF Clone (Origene Technologies) was used. Cells were transfected with Lipofectamine 3000 (Invitrogen) as described by the vendor. TACR3-3’UTR-containing plasmids or empty vectors were combined in a 1:1 mixture with RBMX encoding plasmid or empty vector.

#### 4.5 Animal experiments

SAMR1 and SAMP8 mice (Envigo) were housed under standard condition with free access to water and food under control of the TARC (JGU Mainz). Animals (aged 35-42 weeks) were sacrificed by inhalation anesthesia with Isoflurane and decapitated. Brains were dissected and washed with PBS.

##### Brain slice culture and transfection

Slices of 200 µm were produced (sagittal, McIlwain Tissue Chopper) and the four slices in proximity of the hemispheric cleft (omitting the cerebellum) were used for investigations. Culture plates (12 well) with TC inserts (PET, transparent, pore size: 0,4µm, Sarstedt) were prepared with 1 ml pre-warmed culture medium (B27–2% L-glutamine – 2mM ad. Neurobasal A medium) in each compartment. Slices were incubated at 37°C, 5% CO₂ and 95% humidity in culture medium for 24h. 200ng/µl of endotoxin-free plasmid were diluted 1:50 with Opti-MEM in a volume of 125 µl. Lipofectamine 3000 was diluted ad 1µl/0.033ml Opti-MEM. 125 µl of the reagent mixture were added to the DNA-mixture and incubated at RT for 10min. Half of the supernatant was removed from both compartments of the Transwell system and 250 µl of reagent/DNA-Opti-MEM mixture added to each brain slice. After 30min of incubation 500 µl fresh medium were added to both compartments and slices incubated for 48h. A tissue punch from the area of the hippocampus was extracted by using a 2 mm biopsy needle and lysed in KPi buffer at 10 µl/mg tissue using a tissue lyzer with pre-chilled stainless-steel beads (Qiagen). Supernatans were collected after centrifugation at 10.000 g, 4 °C, 5 min and used for luciferase assay with a sample volume of 10 µl as described for SIM-A9 cells.

##### Western blotting

Tissue specimen were weighed and homogenized in 10 µl homogenization buffer (150 mM NaCl, 1 % Triton X-100, 0.1 %SDS, 50 mM Tris-HCl, per 10mL one cOmplete tablet, Mini EDTA-free, EASYpack) /mg wet weight using a tissue lyzer with pre-chilled stainless-steel beads (Qiagen). Tissue weights were used for analyses comparing SAMR1 and SAMP8.

Supernatants were collected after centrifugation at 10.000 g, 4 °C, 5 min and aliquoted for subsequent analyses and protein quantitation. Forty µg of protein were subjected per lane of a 10% polyacrylamide gel and after gel electrophoresis transferred on nitrocellulose membrane (Amersham). Regular transfer and sample loading was proven by PonceauS red staining. Membranes were blocked in 5% (w/v) milk powder in TBS-Tween20 buffer for 1h at RT. Primary antibodies were as follows: rabbit anti-TACR3-antibody (Biorbyt Cambridge, UK), rabbit anti-RBMX-antibody (Origene Rockland (MD), USA). Secondary antibodies (HRP-labelled) were from ThermoFisherScientific. Primary antibodies were administered over night at 4°C, secondary antibodies for 1h at RT under constant shaking. Detection of signals was performed using the SuperSignalTM West Femto kit (ThermoFisherScientific) and a Stella camera (Raytest).

##### AChE activity assay

One µl of fresh tissue supernatant (see above) was added into a well of a transparent 96 well plate. To this volume, 99 µl of KPi-buffer containing 5mM ACTI and 5 mM DTNB (both Sigma-Aldrich) were added to allow enzyme activity measurement following Elman (Ref). Signals were detected in a Sunrise microplate reader (Tecan) at 412 nm for 45 min with one measurement per minute. Steepness was read from the linear range and calculated per min.

### 5. Statistical analysis

#### 5.1 General approach

Statistical significance was set at α = 0.05. Continuous variables were z-standardized except for interaction terms, voxelwise analyses, and outcomes measured in interpretable units (e.g., years for PhenoAge). For mixed-effects models, p-values were calculated using Satterthwaite’s approximation (Kuznetsova, Brockhoff and Christensen, 2017). Multiple comparisons in voxelwise analyses were controlled using false discovery rate (FDR) correction at FDR = 0.05.

#### 5.2 Neuroimaging analyses

##### 5.2.1 Voxelwise gray matter volume

To test for voxelwise GMV differences in the hippocampus and basal forebrain for rs2765 genotypes in the AgeGain and ADNI cohorts, we used ANCOVA as implemented in the TFCE toolbox for SPM12. GMV was set as dependent variable, while dichotomous rs2765 was set as predictor and age and total intracranial volume (TIV) as covariates. The analysis was masked using the ROIs of the hippocampus and basal forebrain of the neuromorphometrics atlas as provided with CAT12. For a more exploratory approach, we repeated the analysis for the whole brain without prior ROI masking.

##### 5.2.2 Hippocampal volume (ROI-based)

To investigate hippocampal volume differences based on automatic segmentation, we used multiple regression with hippocampal volume as dependent variable, dichotomous rs2765 as predictor and age and TIV as covariates for the AgeGain cohort. To accommodate longitudinal assessments in the ADNI cohort, we estimated the same model using a mixed-effects approach with an additional random intercept per subject.

###### AgeGain

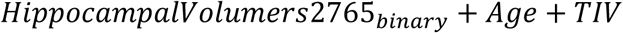

###### ADNI

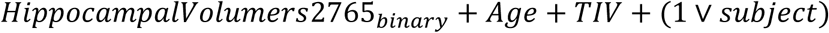

##### 5.2.3 White matter integrity

To exploratively investigate potential voxelwise FA differences along the WM skeleton for rs2765 genotype, we used ANCOVA in the TBSS toolbox of FSL with TFCE. FA values were set as dependent variable, while dichotomous rs2765 was set as predictor and age as covariate.

P-values for all voxelwise analyses were FDR-adjusted for error inflation at FDR = 0.05.

All statistical models subsequently discussed in this section were estimated using R with a significance threshold of α = 0.05.

##### 5.2.4 Brain age estimation

To test whether rs2765 genotype was associated with brain age, we performed a two-sided Mann-Whitney test on the BrainAge Gap between WT and rs2765 variant carriers.

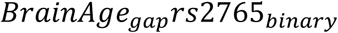

##### 5.2.5 TACR3 expression-structure relationship

To investigate the functional specificity of rs2765 effects on brain structure, we integrated GMV data from the AgeGain cohort with *TACR3* expression data from the Allen Human Brain Atlas (Hawrylycz *et al*., 2012). GMV data corresponding to the expression data were extracted from the individual CAT12 (Gaser *et al*., 2024) modulated and non-linearly spatially normalized GMV images of the AgeGain cohort as the (jacobian modulated) voxel volume at the MNI coordinates corresponding to the expression data points. The lower 20% quantile of these GMV data were removed in order to lower the effect of potential spatial normalization inaccuracies. Based on the mapping of expression measurements to brain regions provided in the Allen data, we classified brain regions as either “HippoBF” (hippocampus/basal forebrain, where we found rs2765 GMV effects) or “Rest” (all other brain regions).

*TACR3* expression values were obtained from the Allen Human Brain Atlas as described in Section 3.4.

We fitted mixed-effects models with local GMV as outcome:

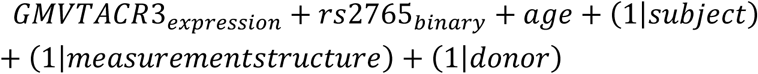

This analysis was performed separately for HippoBF regions (n=12 regions) and Rest regions (n=173 regions) to test our hypothesis that *TACR3* expression-mediated effects would be specific to regions showing rs2765-related structural differences. For validation, the model was also estimated on the whole data set but with an additional interaction term of rs2765 with the classification of Rest vs. HippoBF. Mixed-effects modeling accounted for the nested structure of multiple brain regions within individual donor brains.

#### 5.3 Cognitive performance analyses

##### 5.3.1 Cross-sectional cognition (AgeGain)

To examine whether rs2765 genotype effects on verbal learning and memory (VLMT7) were moderated by age, we fitted an ordinary least squares (OLS) regression model:

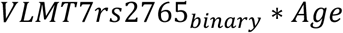

With:

- *rs*2765_*binary*_: Binary indicator of rs2765 genotype (0=WT, 1=non-WT)
- *Age*: Representing age at assessment

This model specification parallels our approach in the ADNI cohort to maintain methodological consistency. Statistical significance was assessed at α = 0.05.

##### 5.3.2 Longitudinal cognitive decline (ADNI)

To investigate a potential effect of rs2765 on cognition and cognitive decline in healthy aging, we selected available longitudinal cognitive data of clinically healthy subjects (at baseline and afterwards) that were classified as amyloid negative during their enrollment in the study based on amyloid PET assessments. We estimated a mixed-effects model on the data with ADAS-cog as dependent variable, age at assessment and dichotomous rs2765 genotype as well as their interaction term. A random intercept per subject was also included.

Furthermore, to investigate, whether the effect of rs2765 on cognition was moderated by developing or present Alzheimer’s disease (AD) pathology, we estimated a modified model on the full ADNI cohort, i.e. subjects with any cognitive status (cognitively normal, mild cognitive impairment, dementia) and available longitudinal cognitive assessments, as well as amyloid PET and rs2765 genotyping. The model was adapted by adding a dichotomous variable encoding, whether the subject had developing or present amyloidosis at any point during enrollment as indicated by at least one amyloid positive PET-assessment, and all pairwise and second order interaction terms of this variable with age at assessment and rs2765 genotype.

###### Amyloid negative ADNI samples (Cognitively healthy aging model)

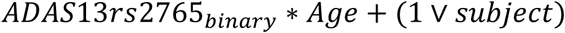

###### Full ADNI cohort

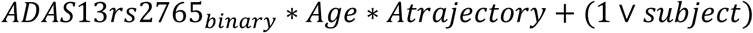

With:

- *AD* – *trajectory*: Developing or present amyloidosis ever during enrollment

#### 5.4 Systemic aging biomarkers

##### 5.4.1 Epigenetic age analysis (PhenoAge)

For epigenetic clock analyses, beta values were extracted immediately after ChAMP preprocessing (pre-filtering for differential methylation) to preserve maximum coverage of CpG sites required for clock algorithms. Epigenetic age was calculated using the methylclock (Pelegí-Sisó *et al*., 2021) R package (version 1.4.0), with verification that all 513 CpG sites required for the PhenoAge clock (Levine *et al*., 2018) were present in our dataset.

The rs2765 variant was analyzed using complementary gene-dose (Figure 3A,C) and binary classification (Figure 3B,D) approaches. Participants were classified as wild-type (WT:GG), heterozygous (Het:AG), or homozygous variant carriers (Hom:AA), with additional binary classification as WT versus non-WT (Het + Hom combined). Group differences were initially assessed using Kruskal-Wallis and Mann-Whitney U tests (Figure 3A,B).

Two complementary ordinary least squares (OLS) regression models were fitted to examine the association between rs2765 genotype and PhenoAge (Figure 3B,D) while controlling for potential confounders. Sex was included as a covariate in PhenoAge analyses due to well-established sex differences in epigenetic aging rates and DNA methylation patterns (Masser *et al*., 2017; Yusipov *et al*., 2020) :

###### Gene-dose-effect model

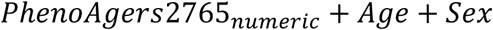

###### Binary genotype model

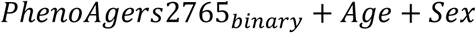

With:

- *rs*2765_*numeric*_: Linear encoding (0=WT, 1=Het, 2=Hom)
- Sex: Binary indicator (0=female, 1=male)

For the gene-dose model, β₁ represents the expected change in PhenoAge (years) per additional variant allele. For the binary model, β₁ represents the expected difference in PhenoAge between non-WT and WT carriers.

All analyses were conducted in Python using scipy.stats for non-parametric tests and statsmodels for regression modeling. Statistical significance was set at α = 0.05, with 95% confidence intervals reported for all effect estimates.

##### 5.4.2 Differential methylation analysis

Differentially methylated positions (DMPs) were identified using the champ.DMP function from the ChAMP package, comparing methylation patterns between WT and non- WT genotypes for rs2765. DMPs were identified using an adjusted p-value threshold of 0.05 (Benjamini-Hochberg correction) to control for multiple testing. All differentially methylated CPGs are available in Supplementary Table 2 based on a p_adj_ ≤ 0.1 basis for exploratory analyses.

##### 5.4.3 Functional enrichment analysis of differential methylation

To characterize the biological significance of differentially methylated CpG sites associated with rs2765 genotype, we performed functional enrichment analysis using hypergeometric testing designed to address inherent bias in gene set analysis of methylation array data(Geeleher *et al*., 2013) .

Rather than directly mapping CpGs to genes then pathways, our approach evaluated CpG-pathway associations by considering the number of CpGs linked to each pathway relative to background distribution. For each functional term, we calculated: (1) total CpG sites globally (N) and specifically associated with all genes in that given term (K), as derived from Illumina EPIC Manifest v1.0B4, (2) number of found differentially methylated CpGs in total (n) and as associated with genes in that given term (k), and (3) finally, enrichment significance using hypergeometric testing with Benjamini-Hochberg multiple testing correction.

All significant terms arising from the pathway enrichment analysis using Gene Ontology (Ashburner *et al*., 2000), KEGG (Kanehisa and Goto, 2000), Reactome (Croft *et al*., 2011), BioCarta Pathways (Adriaens *et al*., 2008), Wikipathways (Agrawal *et al*., 2024), and Pathway Interaction Database (Schaefer *et al*., 2009) are available in Supplementary Table 3 for comprehensive evaluation.

For visual presentation in Figure 3C, we selected 18 representative pathways from the 228 significantly enriched pathways (>5 CpGs, adjusted p < 0.05) to illustrate key mechanistic domains spanning direct brain effects, transcriptional reprogramming, structural networks, and systemic integration. Selection prioritized statistical significance, mechanistic relevance to established NK3-R functions, and representation across functional domains. We acknowledge that highly significant pathways may represent downstream transcriptional cascades rather than direct NK3-R targets. Complete results for all 228 pathways are provided in Supplementary Table 3.

#### 5.5 Experiments based on murine cells and tissue

##### Luciferase assays

Time-course analysis (Figure 5E): Unpaired two-sided Student’s t-tests comparing WT vs variant reporter activity at 48h and 72h timepoints.

Co-transfection experiments (Figure 5F) and brain slice cultures (Figure 5G): One-way ANOVA with Sidak’s post-test for multiple comparisons.

##### Western blot quantification

Between-strain comparisons (SAMR1 vs SAMP8) were performed using unpaired two-sided Student’s t-tests (Figure 6B-C).

##### Protein correlations

Pearson correlations between NK3-R and RBMX (Figure 6D), hippocampal weight (Figure 6E), and AChE activity (Figure 6F) were tested using one-sided tests based on directional hypotheses (positive for D and E; negative for F).

All analyses conducted in GraphPad Prism version 10 with statistical significance set at α = 0.05.

## Supporting information

Supplementary Table 1 - Sample sizes

Supplementary Table 2 - Statistical model summaries

Supplementary Table 3 - Differentially methylated CpGs

Supplementary Table 4 - Pathway enrichment results

Supplementary Figure 1 - UpSet plot of data availability

## Acknowledgement

The AgeGain study was funded by the German Federal Ministry of Education and Research (BMBF) (grant number: 01GQ1425A). In addition, the project was funded by the Leibniz Research Alliance “Resilient Ageing” (Project number: LFV-2021-2-LIR). The German Center for Mental Health (DZPG) is funded by the Federal Ministry of Education and Research (BMBF) German Center for Mental Health [DZPG, BMBF grant 01 EE2305C]; “Science of Healthy Ageing Research Programme” (SHARP) initiative and the Centre of Healthy Ageing (CHA) funded by the Ministry of Science and Health (MWG) of Rhineland-Palatinate, Germany. The authors would also like to thank Rolf Postina (JGU Mainz) for providing the modified pUC19 plasmid.

Data collection and sharing for this project was funded by the Alzheimer’s Disease Neuroimaging Initiative (ADNI) (National Institutes of Health Grant U01 AG024904) and DOD ADNI (Department of Defense award number W81XWH-12-2-0012). ADNI is funded by the National Institute on Aging, the National Institute of Biomedical Imaging and Bioengineering, and through generous contributions from the following: AbbVie, Alzheimer’s Association; Alzheimer’s Drug Discovery Foundation; Araclon Biotech; BioClinica, Inc.; Biogen; Bristol-Myers Squibb Company; CereSpir, Inc.; Cogstate; Eisai Inc.; Elan Pharmaceuticals, Inc.; Eli Lilly and Company; EuroImmun; F. Hoffmann-La Roche Ltd and its affiliated company Genentech, Inc.; Fujirebio; GE Healthcare; IXICO Ltd.; Janssen Alzheimer Immunotherapy Research & Development, LLC.; Johnson & Johnson Pharmaceutical Research & Development LLC.; Lumosity; Lundbeck; Merck & Co., Inc.; Meso Scale Diagnostics, LLC.; NeuroRx Research; Neurotrack Technologies; Novartis Pharmaceuticals Corporation; Pfizer Inc.; Piramal Imaging; Servier; Takeda Pharmaceutical Company; and Transition Therapeutics. The Canadian Institutes of Health Research is providing funds to support ADNI clinical sites in Canada. Private sector contributions are facilitated by the Foundation for the National Institutes of Health (www.fnih.org). The grantee organization is the Northern California Institute for Research and Education, and the study is coordinated by the Alzheimer’s Therapeutic Research Institute at the University of Southern California. ADNI data are disseminated by the Laboratory for Neuro Imaging at the University of Southern California.

## Author contributions

N.R. and F.F. contributed equally as co-first authors. K.E. and O.T. contributed equally as co-senior authors.

N.R.: Conceptualization, Data curation, Formal analysis, Investigation, Methodology, Project administration, Software, Validation, Visualization, Writing – original draft, Writing – review & editing. F.F.: Conceptualization, Data curation, Formal analysis, Investigation, Methodology, Project administration, Software, Validation, Visualization, Writing – original draft, Writing – review & editing. K.E.: Formal analysis, Investigation, Methodology, Resources, Supervision, Validation, Writing – original draft, Writing – review & editing. R.S.S.: Investigation, Validation. J.G.: Investigation, Validation. L.S.: Investigation, Validation. T.S.: Data curation, Project administration. A.P.: Investigation, Methodology. J.W.: Supervision, Writing – review & editing. K.K.: Conceptualization, Funding acquisition, Supervision, Writing – review & editing. B.B.: Conceptualization, Funding acquisition, Supervision, Writing – review & editing. A.M.: Conceptualization, Funding acquisition, Supervision, Writing – review & editing. H.B.: Conceptualization, Funding acquisition, Supervision, Writing – review & editing. A.D.: Conceptualization, Funding acquisition, Supervision, Writing – review & editing. S.T.: Conceptualization, Funding acquisition, Supervision, Writing – review & editing. A.F.: Conceptualization, Funding acquisition, Supervision, Writing – review & editing. S.H.: Methodology, Writing – review & editing. H.M.: Methodology, Writing – review & editing. O.T.: Conceptualization, Funding acquisition, Supervision, Writing – review & editing.

## Data availability

The AgeGain dataset contains sensitive genetic and neuroimaging data from participants who did not provide consent for public data sharing. Accordingly, individual-level AgeGain data cannot be made publicly available. Requests for access to aggregated or summary-level data may be directed to the corresponding author and will be considered on a case-by-case basis in accordance with the study’s ethical approvals and data protection regulations.

ADNI data used in this study are publicly available to qualified researchers through the standard ADNI application process at http://adni.loni.usc.edu. Researchers must complete a data use agreement before accessing ADNI datasets.

Allen Human Brain Atlas expression data are publicly available at https://human.brain-map.org/.

Summary statistics, analysis code, and processed results supporting the findings of this study are available from the corresponding author upon reasonable request.

## Supplement

- Supp. Table 1 – Sample Sizes
- Supp. Table 2 - All statistical models used in xlsx file
- Supp. Table 3 - ChAMP DMP data (Table)
- Supp. Table 4 - All Pathway results (Table)
- Supp. Figure 1 – Upsetplot of sample sizes

## References

1000 Genomes Project Consortium et al. (2015) “A global reference for human genetic variation,” Nature, 526(7571), pp. 68–74. Available at: 10.1038/nature15393.

Adriaens, M.E. et al. (2008) “The public road to high-quality curated biological pathways,” Drug discovery today, 13(0), pp. 856–862. Available at: 10.1016/j.drudis.2008.06.013.

Agrawal, A. et al. (2024) “WikiPathways 2024: next generation pathway database,” Nucleic Acids Research, 52(D1), pp. D679–D689. Available at: 10.1093/nar/gkad960.

Andersson, J.L.R. and Sotiropoulos, S.N. (2016) “An integrated approach to correction for off-resonance effects and subject movement in diffusion MR imaging,” NeuroImage, 125, pp. 1063–1078. Available at: 10.1016/j.neuroimage.2015.10.019.

Ashburner, J. et al. (2014) “SPM12 manual,” Wellcome Trust Centre for Neuroimaging, London, UK, 2464(4), p. 53.

Ashburner, M. et al. (2000) “Gene ontology: tool for the unification of biology. The Gene Ontology Consortium,” Nature Genetics, 25(1), pp. 25–29. Available at: 10.1038/75556.

Boskovic, Z. et al. (2019) “Regulation of cholinergic basal forebrain development, connectivity, and function by neurotrophin receptors,” Neuronal Signaling, 3(1), p. NS20180066. Available at: 10.1042/NS20180066.

Boyle, C.A. et al. (2022) “Ionic signalling mechanisms involved in neurokinin-3 receptor-mediated augmentation of fear-potentiated startle response in the basolateral amygdala,” The Journal of Physiology, 600(19), pp. 4325–4345. Available at: 10.1113/JP283433.

Cardenas, A. et al. (2022) “Epigenome-wide association study and epigenetic age acceleration associated with cigarette smoking among Costa Rican adults,” Scientific Reports, 12(1), p. 4277. Available at: 10.1038/s41598-022-08160-w.

Cheng, X., Zhou, W. and Zhang, Y. (2014) “The behavioral, pathological and therapeutic features of the senescence-accelerated mouse prone 8 strain as an Alzheimer’s disease animal model,” Ageing Research Reviews, 13, pp. 13–37. Available at: 10.1016/j.arr.2013.10.002.

Chételat, G. et al. (2010) “Larger temporal volume in elderly with high versus low beta-amyloid deposition,” Brain, 133(11), pp. 3349–3358. Available at: 10.1093/brain/awq187.

Cook, K.B. et al. (2011) “RBPDB: a database of RNA-binding specificities,” Nucleic Acids Research, 39(suppl_1), pp. D301–D308. Available at: 10.1093/nar/gkq1069.

Cordero, A.I.H. et al. (2024) “Cannabis smoking is associated with advanced epigenetic age,” European Respiratory Journal, 63(5). Available at: 10.1183/13993003.00458-2024.

Cosentino, S. et al. (2008) “APOE ε4 allele predicts faster cognitive decline in mild Alzheimer’s disease,” Neurology, 70(19 Pt 2), pp. 1842–1849. Available at: 10.1212/01.wnl.0000304038.37421.cc.

Croft, D. et al. (2011) “Reactome: a database of reactions, pathways and biological processes,” Nucleic Acids Research, 39(Database issue), pp. D691–D697. Available at: 10.1093/nar/gkq1018.

Cui, F. et al. (2024) “Early-life exposure to tobacco, genetic susceptibility, and accelerated biological aging in adulthood,” Science Advances, 10(18), p. eadl3747. Available at: 10.1126/sciadv.adl3747.

dbSNP rs2765 (2025). Available at: https://www.ncbi.nlm.nih.gov/snp/rs2765 (Accessed: November 11, 2025).

Dong, Q. et al. (2018) “Regulatory RNA binding proteins contribute to the transcriptome-wide splicing alterations in human cellular senescence,” Aging, 10(6), pp. 1489–1505. Available at: 10.18632/aging.101485.

Dyck, C.H. van et al. (2023) “Lecanemab in Early Alzheimer’s Disease,” New England Journal of Medicine, 388(1), pp. 9–21. Available at: 10.1056/NEJMoa2212948.

Elliott, D.J. et al. (2019) “RBMX family proteins connect the fields of nuclear RNA processing, disease and sex chromosome biology,” The International Journal of Biochemistry & Cell Biology, 108, pp. 1–6. Available at: 10.1016/j.biocel.2018.12.014.

Ferrucci, L. and Kuchel, G.A. (2021) “Heterogeneity of Aging: Individual Risk Factors, Mechanisms, Patient Priorities, and Outcomes,” Journal of the American Geriatrics Society, 69(3), pp. 610–612. Available at: 10.1111/jgs.17011.

Fischer, F.U. et al. (2025) “Cognitive training gain transfer in cognitively healthy aging: per protocol results of the German AgeGain study,” Frontiers in Aging Neuroscience, 17. Available at: 10.3389/fnagi.2025.1587395.

Fitzgerald, K.N. et al. (2023) “Potential reversal of biological age in women following an 8-week methylation-supportive diet and lifestyle program: a case series,” Aging, 15(6), pp. 1833–1839. Available at: 10.18632/aging.204602.

Gaser, C. et al. (2024) “CAT: a computational anatomy toolbox for the analysis of structural MRI data,” GigaScience, 13, p. giae049. Available at: 10.1093/gigascience/giae049.

Geeleher, P. et al. (2013) “Gene-set analysis is severely biased when applied to genome-wide methylation data,” *Bioinformatics (Oxford*, England), 29(15), pp. 1851–1857. Available at: 10.1093/bioinformatics/btt311.

Griesemer, D. et al. (2021) “Genome-wide functional screen of 3’UTR variants uncovers causal variants for human disease and evolution,” Cell, 184(20), pp. 5247–5260.e19. Available at: 10.1016/j.cell.2021.08.025.

Hartig, M. et al. (2014) “UCSF freesurfer methods,” ADNI Alzheimers Disease Neuroimaging Initiative: San Francisco, CA, USA, 5.

Hawrylycz, M.J. et al. (2012) “An anatomically comprehensive atlas of the adult human brain transcriptome,” Nature, 489(7416), pp. 391–399. Available at: 10.1038/nature11405.

Huang, H.-Z. et al. (2024) “Senktide blocks aberrant RTN3 interactome to retard memory decline and tau pathology in social isolated Alzheimer’s disease mice,” Protein & Cell, 15(4), pp. 261–284. Available at: 10.1093/procel/pwad056.

Johnson, K.A. et al. (2023) “Efficacy and Safety of Fezolinetant in Moderate to Severe Vasomotor Symptoms Associated With Menopause: A Phase 3 RCT,” The Journal of Clinical Endocrinology and Metabolism, 108(8), pp. 1981–1997. Available at: 10.1210/clinem/dgad058.

Kalc, P. et al. (2024) “BrainAGE: Revisited and reframed machine learning workflow,” Human Brain Mapping, 45(3), p. e26632. Available at: 10.1002/hbm.26632.

Kanehisa, M. and Goto, S. (2000) “KEGG: kyoto encyclopedia of genes and genomes,” Nucleic Acids Research, 28(1), pp. 27–30. Available at: 10.1093/nar/28.1.27.

Ke, T.-M., Lophatananon, A. and Muir, K.R. (2024) “Exploring the Relationships between Lifestyle Patterns and Epigenetic Biological Age Measures in Men,” Biomedicines, 12(9), p. 1985. Available at: 10.3390/biomedicines12091985.

Koca, R.O. et al. (2023) “How does neurokinin 3 receptor agonism affect pathological and cognitive impairments in an Alzheimer’s disease-like rat model?,” Amino Acids, 55(4), pp. 481–498. Available at: 10.1007/s00726-023-03241-0.

Koca, R.O. et al. (2024) “Are the promnestic effects of neurokinin 3 receptor mediated by hippocampal neurogenesis in a Aβ-induced rat model of Alzheimer’s disease?,” International Journal of Developmental Neuroscience: The Official Journal of the International Society for Developmental Neuroscience, 84(7), pp. 688–703. Available at: 10.1002/jdn.10362.

Kueper, J.K., Speechley, M. and Montero-Odasso, M. (2018) “The Alzheimer’s Disease Assessment Scale–Cognitive Subscale (ADAS-Cog): Modifications and Responsiveness in Pre-Dementia Populations. A Narrative Review,” Journal of Alzheimer’s Disease, 63(2), pp. 423–444. Available at: 10.3233/JAD-170991.

Kuznetsova, A., Brockhoff, P.B. and Christensen, R.H.B. (2017) “lmerTest Package: Tests in Linear Mixed Effects Models,” Journal of Statistical Software, 82, pp. 1–26. Available at: 10.18637/jss.v082.i13.

Le Stanc, L. et al. (2024) “Cognitive reserve involves decision making and is associated with left parietal and hippocampal hypertrophy in neurodegeneration,” Communications Biology, 7(1), p. 741. Available at: 10.1038/s42003-024-06416-x.

Lederman, S. et al. (2023) “Fezolinetant for treatment of moderate-to-severe vasomotor symptoms associated with menopause (SKYLIGHT 1): a phase 3 randomised controlled study,” *Lancet (London*, England), 401(10382), pp. 1091–1102. Available at: 10.1016/S0140-6736(23)00085-5.

Levine, M.E. et al. (2018) “An epigenetic biomarker of aging for lifespan and healthspan,” Aging (Albany NY), 10(4), pp. 573–591. Available at: 10.18632/aging.101414.

Lin, J.-R. et al. (2021) “Rare genetic coding variants associated with human longevity and protection against age-related diseases,” Nature Aging, 1(9), pp. 783–794. Available at: 10.1038/s43587-021-00108-5.

Liu, B., Liu, J. and Shi, J.-S. (2020) “SAMP8 Mice as a Model of Age-Related Cognition Decline with Underlying Mechanisms in Alzheimer’s Disease,” Journal of Alzheimer’s disease: JAD, 75(2), pp. 385–395. Available at: 10.3233/JAD-200063.

Masser, D.R. et al. (2017) “Sexually divergent DNA methylation patterns with hippocampal aging,” Aging Cell, 16(6), pp. 1342–1352. Available at: 10.1111/acel.12681.

Menegaz de Almeida, A., et al. (2025) “Fezolinetant and Elinzanetant Therapy for Menopausal Women Experiencing Vasomotor Symptoms: A Systematic Review and Meta-analysis,” Obstetrics and Gynecology, 145(3), pp. 253–261. Available at: 10.1097/AOG.0000000000005812.

Mohs, R.C., Rosen, W.G. and Davis, K.L. (1983) “The Alzheimer’s disease assessment scale: an instrument for assessing treatment efficacy,” Psychopharmacology Bulletin, 19(3), pp. 448–450.

Morales Berstein, F., et al. (2022) “Assessing the causal role of epigenetic clocks in the development of multiple cancers: a Mendelian randomization study,” eLife. Edited by M.M. Rovers, E. Franco, and D.W. Belsky, 11, p. e75374. Available at: 10.7554/eLife.75374.

Neal-Perry, G. et al. (2023) “Safety of Fezolinetant for Vasomotor Symptoms Associated With Menopause: A Randomized Controlled Trial,” Obstetrics and Gynecology, 141(4), pp. 737–747. Available at: 10.1097/AOG.0000000000005114.

Nestler, E.J. and Russo, S.J. (2024) “Neurobiological basis of stress resilience,” Neuron, 112(12), pp. 1911–1929. Available at: 10.1016/j.neuron.2024.05.001.

Pelegí-Sisó, D. et al. (2021) “methylclock: a Bioconductor package to estimate DNA methylation age,” Bioinformatics, 37(12), pp. 1759–1760. Available at: 10.1093/bioinformatics/btaa825.

Prieto, C. et al. (2021) “Transcriptional control of CBX5 by the RNA binding proteins RBMX and RBMXL1 maintains chromatin state in myeloid leukemia,” Nature Cancer, 2, pp. 741–757. Available at: 10.1038/s43018-021-00220-w.

Ray, N.J. et al. (2015) “Cholinergic basal forebrain structure influences the reconfiguration of white matter connections to support residual memory in mild cognitive impairment,” The Journal of Neuroscience: The Official Journal of the Society for Neuroscience, 35(2), pp. 739–747. Available at: 10.1523/JNEUROSCI.3617-14.2015.

Rohde, S.K. et al. (2025) “Amyloid-Beta Pathology and Cognitive Performance in Centenarians,” JAMA neurology, 82(8), pp. 837–847. Available at: 10.1001/jamaneurol.2025.1734.

Romo, L., Findlay, S.D. and Burge, C.B. (2024) “Regulatory features aid interpretation of 3′UTR variants,” American Journal of Human Genetics, 111(2), pp. 350–363. Available at: 10.1016/j.ajhg.2023.12.017.

Schaefer, C.F. et al. (2009) “PID: the Pathway Interaction Database,” Nucleic Acids Research, 37(Database issue), pp. D674-679. Available at: 10.1093/nar/gkn653.

Schmitz, T.W., Nathan Spreng, R., and Alzheimer’s Disease Neuroimaging Initiative (2016) “Basal forebrain degeneration precedes and predicts the cortical spread of Alzheimer’s pathology,” Nature Communications, 7, p. 13249. Available at: 10.1038/ncomms13249.

Sims, J.R. et al. (2023) “Donanemab in Early Symptomatic Alzheimer Disease,” JAMA, 330(6), pp. 512–527. Available at: 10.1001/jama.2023.13239.

de Souza Silva, M.A., et al. (2013) “Neurokinin3 receptor as a target to predict and improve learning and memory in the aged organism,” Proceedings of the National Academy of Sciences of the United States of America, 110(37), pp. 15097–15102. Available at: 10.1073/pnas.1306884110.

Stern, Y. et al. (2020) “Whitepaper: Defining and investigating cognitive reserve, brain reserve, and brain maintenance,” Alzheimer’s & Dementia: The Journal of the Alzheimer’s Association, 16(9), pp. 1305–1311. Available at: 10.1016/j.jalz.2018.07.219.

Suzuki, K. et al. (2020) “Effect of apolipoprotein E ε4 allele on the progression of cognitive decline in the early stage of Alzheimer’s disease,” Alzheimer’s & Dementia: Translational Research & Clinical Interventions, 6(1), p. e12007. Available at: 10.1002/trc2.12007.

Teipel, S.J. et al. (2015) “Association of a neurokinin 3 receptor polymorphism with the anterior basal forebrain,” Neurobiology of Aging, 36(6), pp. 2060–2067. Available at: 10.1016/j.neurobiolaging.2014.12.031.

Tesi, N. et al. (2024) “Cognitively healthy centenarians are genetically protected against Alzheimer’s disease,” Alzheimer’s & Dementia, 20(6), pp. 3864–3875. Available at: 10.1002/alz.13810.

Tian, Y. et al. (2017) “ChAMP: updated methylation analysis pipeline for Illumina BeadChips,” Bioinformatics, 33(24), pp. 3982–3984. Available at: 10.1093/bioinformatics/btx513.

Tian, Y.E. et al. (2023) “Heterogeneous aging across multiple organ systems and prediction of chronic disease and mortality,” Nature Medicine, 29(5), pp. 1221–1231. Available at: 10.1038/s41591-023-02296-6.

Varesi, A. et al. (2023) “RNA binding proteins in senescence: A potential common linker for age-related diseases?,” Ageing Research Reviews, 88, p. 101958. Available at: 10.1016/j.arr.2023.101958.

Wang, W. et al. (2023) “trRosettaRNA: automated prediction of RNA 3D structure with transformer network,” Nature Communications, 14(1), p. 7266. Available at: 10.1038/s41467-023-42528-4.

Whitehair, D.C. et al. (2010) “Influence of Apolipoprotein E ε4 on rates of cognitive and functional decline in mild cognitive impairment,” Alzheimer’s & dementia : the journal of the Alzheimer’s Association, 6(5), pp. 412–419. Available at: 10.1016/j.jalz.2009.12.003.

Wojtas, M.N. et al. (2024) “Interplay between hippocampal TACR3 and systemic testosterone in regulating anxiety-associated synaptic plasticity,” Molecular Psychiatry, 29(3), pp. 686–703. Available at: 10.1038/s41380-023-02361-z.

Wolf, D. et al. (2014) “Association of basal forebrain volumes and cognition in normal aging,” Neuropsychologia, 53, pp. 54–63. Available at: 10.1016/j.neuropsychologia.2013.11.002.

Wolf, D. et al. (2018) “Mechanisms and modulators of cognitive training gain transfer in cognitively healthy aging: study protocol of the AgeGain study,” Trials, 19(1), p. 337. Available at: 10.1186/s13063-018-2688-2.

Wolf, D., Fischer, F.U., Fellgiebel, A., and Alzheimer’s Disease Neuroimaging Initiative (2019) “A methodological approach to studying resilience mechanisms: demonstration of utility in age and Alzheimer’s disease-related brain pathology,” Brain Imaging and Behavior, 13(1), pp. 162–171. Available at: 10.1007/s11682-018-9870-8.

Wolf, D., Fischer, F.U., Fellgiebel, A., and for the Alzheimer’s Disease Neuroimaging Initiative (2019) “A methodological approach to studying resilience mechanisms: demonstration of utility in age and Alzheimer’s disease-related brain pathology,” Brain Imaging and Behavior, 13(1), pp. 162–171. Available at: 10.1007/s11682-018-9870-8.

Wu, D. et al. (2024) “Dose response of leisure time physical activity and biological aging in type 2 diabetes: a cross sectional study,” Scientific Reports, 14(1), p. 26253. Available at: 10.1038/s41598-024-77359-w.

Yusipov, I. et al. (2020) “Age-related DNA methylation changes are sex-specific: a comprehensive assessment,” Aging (Albany NY), 12(23), pp. 24057–24080. Available at: 10.18632/aging.202251.

Zhang, W.-W., Wang, Y. and Chu, Y.-X. (2020) “Tacr3/NK3R: Beyond Their Roles in Reproduction,” ACS chemical neuroscience, 11(19), pp. 2935–2943. Available at: 10.1021/acschemneuro.0c00421.

Zhang, Y., Parmigiani, G. and Johnson, W.E. (2020) “ComBat-seq: batch effect adjustment for RNA-seq count data,” NAR Genomics and Bioinformatics, 2(3), p. lqaa078. Available at: 10.1093/nargab/lqaa078.

