## Supplementary figures and images for "TACR3 variant confers resilience to aging and Alzheimer’s disease"

### Supplementary Figure 1 - UpSet plot of data availability

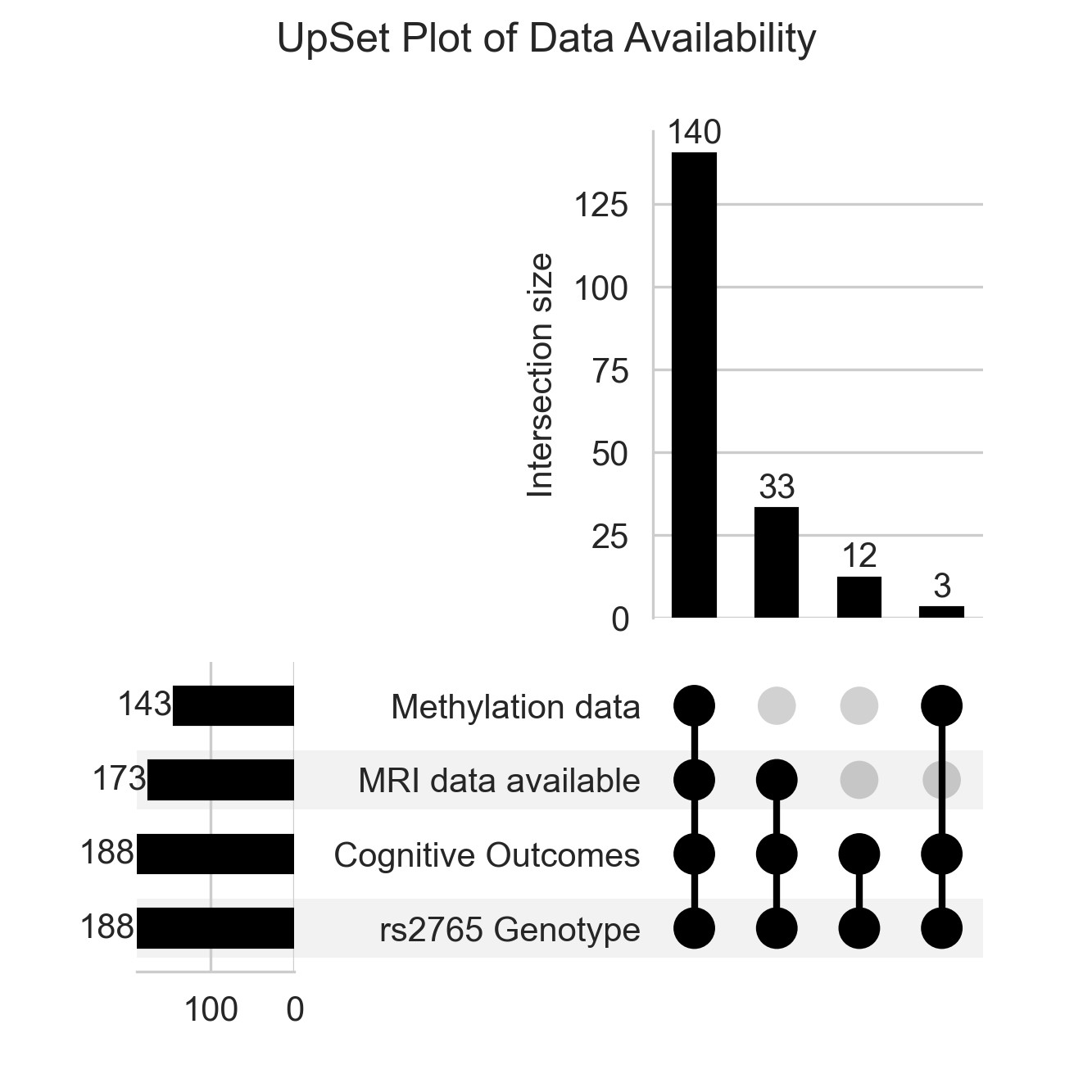
